# Combination of inflammatory and vascular markers in the febrile phase of dengue is associated with more severe outcomes

**DOI:** 10.1101/2021.03.13.21253501

**Authors:** Nguyen Lam Vuong, Phung Khanh Lam, Damien Keng Yen Ming, Huynh Thi Le Duyen, Nguyet Minh Nguyen, Dong Thi Hoai Tam, Duong Thi Hue Kien, Nguyen Van Vinh Chau, Ngoun Chanpheaktra, Lucy Chai See Lum, Ernesto Pleités, Cameron P. Simmons, Kerstin Rosenberger, Thomas Jaenisch, David Bell, Nathalie Acestor, Christine Halleux, Piero L. Olliaro, Bridget A. Wills, Ronald B. Geskus, Sophie Yacoub

**Affiliations:** Oxford University Clinical Research Unit (OUCRU), Ho Chi Minh City, Viet Nam; University of Medicine and Pharmacy at Ho Chi Minh City, Ho Chi Minh City, Viet Nam; Department of Infectious Diseases, Imperial College London, London, UK; Hospital for Tropical Diseases, Ho Chi Minh city, Viet Nam; Angkor Hospital for Children, Siem Reap, Cambodia; University of Malaya Medical Centre, Kuala Lumpur, Malaysia; Hospital Nacional de Niños Benjamin Bloom, San Salvador, El Salvador; Centre for Tropical Medicine and Global health, Nuffield Department of Clinical Medicine, University of Oxford, United Kingdom; Institute for Vector-Borne Disease, Monash University, Clayton, Australia; Section Clinical Tropical Medicine, Department for Infectious Diseases, Heidelberg University Hospital, Heidelberg, Germany; Heidelberg Institute of Global Health (HIGH), Heidelberg University Hospital, Germany; Independent consultant, Issaquah, WA, USA; Consultant, Intellectual Ventures, Global Good Fund, Bellevue, WA, USA; UNICEF/UNDP/World Bank/WHO Special Programme for Research and Training in Tropical Diseases, World Health Organization, Geneva, Switzerland

**Keywords:** Dengue, Biomarkers, Prognostic, IL1Ra, Syndecan

## Abstract

**Background:** Early identification of severe dengue patients is important regarding patient management and resource allocation. We investigated the association of ten biomarkers (VCAM-1, SDC-1, Ang-2, IL-8, IP-10, IL-1RA, sCD163, sTREM-1, ferritin, CRP) with the development of severe/moderate dengue (S/MD).

**Methods:** We performed a nested case-control study from a multi-country study. A total of 281 S/MD and 556 uncomplicated dengue cases were included.

**Results:** On days 1-3 from symptom onset, higher levels of any biomarker increased the risk of developing S/MD. When assessing together, SDC-1 and IL-1RA were stable, while IP-10 changed the association from positive to negative; others showed weaker associations. The best combinations associated with S/MD comprised IL-1RA, Ang-2, IL-8, ferritin, IP-10, and SDC-1 for children, and SDC-1, IL-8, ferritin, sTREM-1, IL-1RA, IP-10, and sCD163 for adults.

**Conclusions:** Our findings assist the development of biomarker panels for clinical use and could improve triage and risk prediction in dengue patients.

**Summary of the main point:** Higher levels of any of VCAM-1, SDC-1, Ang-2, IL-8, IP-10, IL-1RA, sCD163, sTREM-1, ferritin, and CRP on illness days 1-3 increased the risk of developing severe/moderate dengue. The relationships differed between children and adults and some changed when assessed together.

## Introduction

Dengue is the most common arboviral disease to affect humans globally. In 2019, the World Health Organization (WHO) identified dengue as one of the top ten threats to global health;[1] transmission occurs in 129 countries, with an estimated 3.9 billion people being at risk.[2] Over the last two decades, the number of reported cases per year has increased more than eight-fold,[2] and in 2020 the annual number of dengue virus (DENV) infections was estimated to be 105 million, with 51 million cases being clinically apparent.[3] With climate change, increased travel and urbanization, this rise is forecasted to continue over the coming decades.[4,5] Despite the large disease burden, there is still no specific treatment for dengue, and the only licensed vaccine is recommended only in individuals with earlier dengue infection.[6]

In many dengue-endemic settings seasonal epidemics can rapidly overwhelm fragile health systems. Although most symptomatic dengue infections are self-limiting, a small proportion of patients develop complications, most of which manifest at around 4-6 days from symptom onset. Thus, large numbers of patients require regular assessments to identify complications should they arise. The accurate and early identification of such patients, particularly within the first three days of illness in the febrile phase, should allow for appropriate care to be provided and potentially increase health system effectiveness. Although the 2009 WHO dengue guidelines set out specific warning signs for use in patient triage, utility of these guidelines at identifying those at risk for complications remains limited.[7]

The pathogenesis of dengue involves a complex interplay between viral factors and the host response. It is hypothesized that an excessive immune response acting through inflammatory mediators can lead to the observed manifestations of bleeding, shock and organ dysfunction. Studies have shown that in secondary infections, adaptive immune activation can result in high circulating levels of plasma cytokines and chemokines.[8–10] Binding of viral NS1 protein onto endothelial cells can act in concert with vasoactive substances, cytokines and chemokines, to result in endothelial activation and glycocalyx disruption, and these processes likely underlie the increased vascular permeability and coagulopathy.[11–13]

The role of blood biomarkers in predicting severe outcomes has been investigated in many studies, but mostly at later time-points or at hospital admission and many of these biomarkers either peak too late in the disease course or have too short a half-life to be clinically useful.[14–25] Acknowledging these characteristics, we selected ten candidate biomarkers from the vascular, immunological, and inflammatory pathways with good evidence supporting their involvement in the pathogenesis of dengue infection – focusing on those likely to be increased early in the disease course. We included vascular cell adhesion molecule-1 (VCAM-1), syndecan-1 (SDC-1), and angiopoietin-2 (Ang-2) because they represent endothelial activation and glycocalyx integrity.[26–29] For markers of immune activation, we measured interleukin-8 (IL-8) and interferon gamma-induced protein-10 (IP-10) as these are associated with disease severity,[22,30,31] and IL-1 receptor antagonist (IL-1RA), soluble cluster of differentiation 163 (sCD163), and soluble triggering receptor expressed on myeloid cells-1 (sTREM-1) as these are activation markers of monocytes and macrophages, the major targets for dengue replication.[14,21,23] For markers of general inflammation we included ferritin and C-reactive protein (CRP).[21,32-35]

The aims of this study were: (1) to investigate the association of these ten biomarkers with development of more severe dengue outcomes, (2) to find the best combination of biomarkers associated with more severe dengue outcomes. The results of the second aim could help in developing multiplex panels for use in outpatient settings to rapidly identify patients who require hospitalization.

## Methods

### Study design

We conducted a nested case-control study using the samples and clinical information from a large multi-country observational study named “Clinical evaluation of dengue and identification of risk factors for severe disease” (IDAMS study, NCT01550016).[36] The IDAMS study and the blood sample analysis were approved by the Scientific and Ethics Committees of all study sites and by the Oxford Tropical Research Ethics Committee. There were 7,428 participants in eight countries across Asia and Latin America enrolled in the IDAMS study. Patients were eligible for inclusion if they were aged five years or older, had fever or history of fever for less than 72 hours, and had symptoms consistent with dengue, with no features strongly suggestive of another disease. Participants were followed daily with a standard schedule of clinical examination and blood samples. Individual management (including hospitalization) was in accordance with routine practice at each study site. All diagnostic samples were processed and stored following specific protocols, and later transferred to designated sites for diagnostic testing in order to ensure consistency. Laboratory-confirmed dengue was defined by a positive reverse transcriptase polymerase chain reaction (RT-PCR) or a positive NS1 enzyme-linked immunosorbent assay (ELISA) result. Immune status was classified based on capture IgG results on paired samples. A probable primary infection was defined by two negative IgG results on two consecutive specimens taken at least two days apart, with at least one specimen obtained during the convalescent phase (after illness day 5). A probable secondary infection was defined by a positive IgG result identified during either or both the febrile and convalescent phases. In all other cases with the absence of suitable specimens at the appropriate time points immune status was classified as inconclusive. Each participant was given an overall severity grade (severe, moderate, or uncomplicated dengue), using all available information and a grading system in line with current guidelines and recommendations to classify clinical endpoints in dengue clinical trials.[37]

### Study population

Of the 2,694 laboratory-confirmed dengue cases in the IDAMS study, 38 and 266 cases were classified as severe and moderate dengue respectively. For this study, we selected all severe and moderate cases from five study sites in four countries (Vietnam, Cambodia, Malaysia, and El Salvador), as residual plasma from these countries’ sample sets was available at the Oxford University Clinical Research Unit (OUCRU) in Ho Chi Minh City, Vietnam. For the control group, we selected patients with uncomplicated dengue with similar geographic and demographic characteristics at a 2:1 ratio. In total 281 cases and 556 controls were included in the analysis (Figure 1).

**Figure 1.**
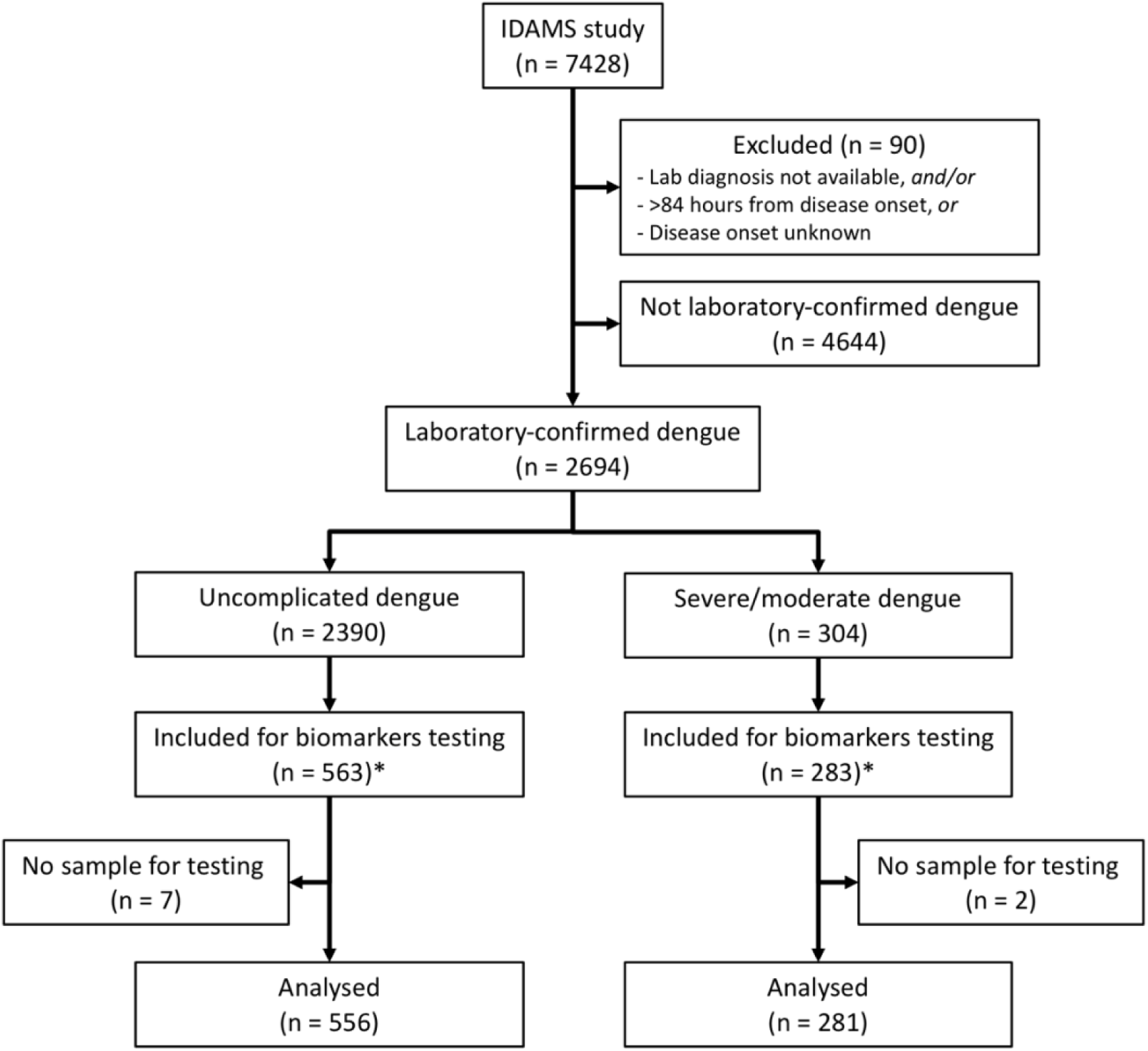
Study flowchart. *The IDAMS study was performed in eight countries across Asia and Latin America. For this study, we selected cases in four countries (Vietnam, Cambodia, Malaysia, and El Salvador) as the blood samples were stored at the laboratory of the Oxford University Clinical Research Unit in Ho Chi Minh City, Vietnam.

### Laboratory evaluation (details in appendix 1)

The biomarkers were measured at two time points: at enrollment (illness day 1-3) and after recovery (day 10-31 post-symptom onset), if available. Eight biomarkers (CRP and ferritin excepted) were combined in a premixed magnetic bead panel (Cat No. LXSAHM; R&D). CRP was measured using a separate commercial magnetic bead panel (Cat. No. HCVD3MAG-67K; EMD Millipore Corporation). These panels were analysed using the Luminex200^TM^ analyzer with the Luminex calibration (Cat. No. LX200-CAL-K25) and verification kits (Cat. No. LX200-CON-K25). Ferritin was measured using the Human Ferritin ELISA kit (Cat. No. ARG80501, Arigo). All tests were done according to the manufacturer’s specifications.

### Study endpoints (details in appendix 2)

The primary endpoint was combined severe and moderate dengue (S/MD), defined by the development of severe or moderate grades of any of the following - plasma leakage, haemorrhage, or organ impairment (including neurologic, hepatic, or cardiac involvement) (Table S1). We combined severe and moderate dengue to form the primary endpoint (S/MD) as severe dengue events were rare; this combined endpoint is relevant to clinical practice since the moderate group is likely to develop complications and therefore may also require medical intervention and hospitalization. We studied three secondary endpoints: severe dengue alone, severe dengue or dengue with warning signs according to the 2009 WHO classification, and hospitalization. These endpoints were selected as they also reflect the disease burden and severity and are generalizable across different settings. The decision to hospitalize was based only on clinical judgement and local guidelines particular to each study site, without use of any biomarker information.

### Statistical analysis (details in appendix 3)

Plasma levels of all biomarkers were transformed to the base-2 logarithm (log-2) before analysis as a right skewed distribution was apparent. We used a logistic regression model for all endpoints. We investigated the non-linear effects of all biomarkers and age on the endpoints, using restricted cubic splines with three knots at the 10^th^, 50^th^, and 90^th^ percentiles.

For the first aim, i.e. to investigate the association of all biomarkers with the primary and secondary endpoints, we performed two different analyses: (1) fitting models for each biomarker separately (‘single models’), and (2) fitting models including all biomarkers together (‘global models’). In the ‘single models’ for a particular biomarker, only that biomarker along with age and their interaction were included, whereas in the ‘global models’ all the biomarkers along with their interactions with age were included. Results are reported as odds ratio (OR) and presented graphically.

To find the best combination of biomarkers associated with the primary endpoint we built upon the results from the first aim to fit separate models for children and adults (<15 versus ≥15 years of age), as differences were apparent by age. We used variable selection based on the ‘best subset’ approach.[38,39] From an ‘initial model’ including all biomarkers, we determined the best general combination and the best combinations of 2, 3, 4, and 5 biomarkers, based on the Akaike information criterion (AIC). We then performed a bootstrap procedure by taking 1,000 samples and check the robustness (stability) of the selected models.

All analyses were performed using the statistical software R version 3.6.3.

## Results

### Patient characteristics

The majority of the patients were from Vietnam (640 cases, 76%). Median (1^st^, 3^rd^ quartiles) age of the case and control groups were 12 (9, 22) and 16 (10, 24) years. Among the S/MD group, 127 cases (45%) were children and 154 cases (55%) were adults. Male gender was predominant (60% and 54% in the case and control groups respectively). Serotype distribution was similar between the S/MD and control groups, with DENV-1 predominating (42%), particularly in children (48%). Host immune status however differed: there was a higher proportion of secondary infections in the S/MD group compared with controls (78% versus 64%, respectively) and this was consistent in both children and adults. Overall, 38 patients developed severe dengue, most were severe plasma leakage (33/38 cases, 87%) and 29/38 (76%) were children. Most of the moderate dengue cases were plasma leakage and/or hepatic involvement. As expected, hospitalization was more common in the S/MD group (57% versus 31%) (Table 1).

**Table 1.** Summary of clinical data by primary outcome.

|  | All patients |  | Children |  | Adults |  |
| --- | --- | --- | --- | --- | --- | --- |
|  | Uncomplicated dengue (N=556) | Severe/moderate dengue (N=281) | Uncomplicated dengue (N=337) | Severe/moderate dengue (N=127) | Uncomplicated dengue (N=219) | Severe/moderate dengue (N=154) |
| Country, *n* (%) |  |  |  |  |  |  |
| - Cambodia | 39 (7) | 30 (11) | 37 (11) | 29 (23) | 2 (1) | 1 (1) |
| - El Salvador | 23 (4) | 18 (6) | 23 (7) | 18 (14) | 0 (0) | 0 (0) |
| - Malaysia | 58 (10) | 29 (10) | 3 (1) | 1 (1) | 55 (25) | 28 (18) |
| - Vietnam | 436 (78) | 204 (73) | 274 (81) | 79 (62) | 162 (74) | 125 (81) |
| Age (years), median (1<sup>st</sup>, 3<sup>rd</sup> quartiles) | 12 (9, 22) | 16 (10, 24) | 10 (8, 12) | 10 (7, 12) | 26 (20, 34) | 22 (18, 30) |
| Gender male, *n* (%) | 299 (54) | 170 (60) | 173 (51) | 80 (63) | 126 (58) | 90 (58) |
| Illness day at enrolment, *n* (%) |  |  |  |  |  |  |
| - 1 | 91 (16) | 49 (17) | 57 (17) | 25 (20) | 34 (16) | 24 (16) |
| - 2 | 260 (47) | 130 (46) | 156 (46) | 52 (41) | 104 (47) | 78 (51) |
| - 3 | 205 (37) | 102 (36) | 124 (37) | 50 (39) | 81 (37) | 52 (34) |
| Serotype, *n* (%) |  |  |  |  |  |  |
| - DENV-1 | 228 (41) | 121 (43) | 161 (48) | 61 (48) | 67 (31) | 60 (39) |
| - DENV-2 | 74 (13) | 47 (17) | 22 (7) | 16 (13) | 52 (24) | 31 (20) |
| - DENV-3 | 59 (11) | 29 (10) | 43 (13) | 18 (14) | 16 (7) | 11 (7) |
| - DENV-4 | 161 (29) | 70 (25) | 91 (27) | 26 (20) | 70 (32) | 44 (29) |
| - Unknown | 34 (6) | 14 (5) | 20 (6) | 6 (5) | 14 (6) | 8 (5) |
| Immune status, *n* (%) |  |  |  |  |  |  |
| - Probable primary | 124 (22) | 41 (15) | 86 (26) | 15 (12) | 38 (17) | 26 (17) |
| - Probable secondary | 355 (64) | 218 (78) | 202 (60) | 100 (79) | 153 (70) | 118 (77) |
| - Inconclusive | 77 (14) | 22 (8) | 49 (15) | 12 (9) | 28 (13) | 10 (6) |
| Severe dengue*, *n* (%) | 0 (0) | 38 (14) | 0 (0) | 29 (23) | 0 (0) | 9 (6) |
| - Severe plasma leakage |  | 33 (12) |  | 24 (19) |  | 9 (6) |
| + Dengue shock syndrome |  | 25 (9) |  | 18 (14) |  | 7 (5) |
| + Respiratory distress |  | 12 (4) |  | 9 (7) |  | 3 (2) |
| - Severe neurologic involvement |  | 3 (1) |  | 3 (2) |  | 0 (0) |
| - Severe bleeding |  | 2 (1) |  | 2 (2) |  | 0 (0) |
| - Severe other major organ failure |  | 1 (0) |  | 0 (0) |  | 1 (1) |
| - Severe hepatic involvement |  | 0 (0) |  | 0 (0) |  | 0 (0) |
| Moderate dengue*, *n* (%) | 0 (0) | 243 (86) | 0 (0) | 98 (77) | 0 (0) | 145 (94) |
| - Moderate plasma leakage |  | 159 (57) |  | 73 (57) |  | 86 (56) |
| - Moderate hepatic involvement |  | 102 (36) |  | 35 (28) |  | 67 (44) |
| - Moderate bleeding |  | 9 (3) |  | 3 (2) |  | 6 (4) |
| - Moderate other major organ involvement |  | 1 (0) |  | 0 (0) |  | 1 (1) |
| - Moderate neurologic involvement |  | 0 (0) |  | 0 (0) |  | 0 (0) |
| WHO 2009 classification, *n* (%) |  |  |  |  |  |  |
| - Mild dengue | 266 (48) | 49 (18) | 168 (50) | 17 (14) | 98 (45) | 32 (21) |
| - Dengue with warning signs | 288 (52) | 186 (67) | 169 (50) | 81 (65) | 119 (55) | 105 (69) |
| - Severe dengue | 0 (0) | 43 (15) | 0 (0) | 27 (22) | 0 (0) | 16 (10) |
| - Unknown | 2 (0) | 3 (1) | 0 (0) | 2 (2) | 2 (1) | 1 (1) |
| Hospitalization, *n* (%) | 175 (31) | 161 (57) | 127 (38) | 83 (65) | 48 (22) | 78 (51) |

\*Individuals can be included in more than one severe dengue categories
WHO: World Health Organization

### Biomarker levels

On average, the patients who progressed to S/MD had higher levels of the biomarkers in both children and adult patients, both at enrollment and at follow-up (Table 2, Figure 2). The levels of five biomarkers (VCAM-1, IL-8, IP-10, IL-1RA, and CRP) decreased between the two time-points, whereas SDC-1 increased slightly and the other markers showed no clear trends (Figure S1). Moderate-to-strong positive correlations were evident for some markers, in particular IP-10 and IL-1RA, and IP-10 and VCAM-1, both with Spearman’s rank correlation coefficients above 0.6 (Figure S2).

**Figure 2.**
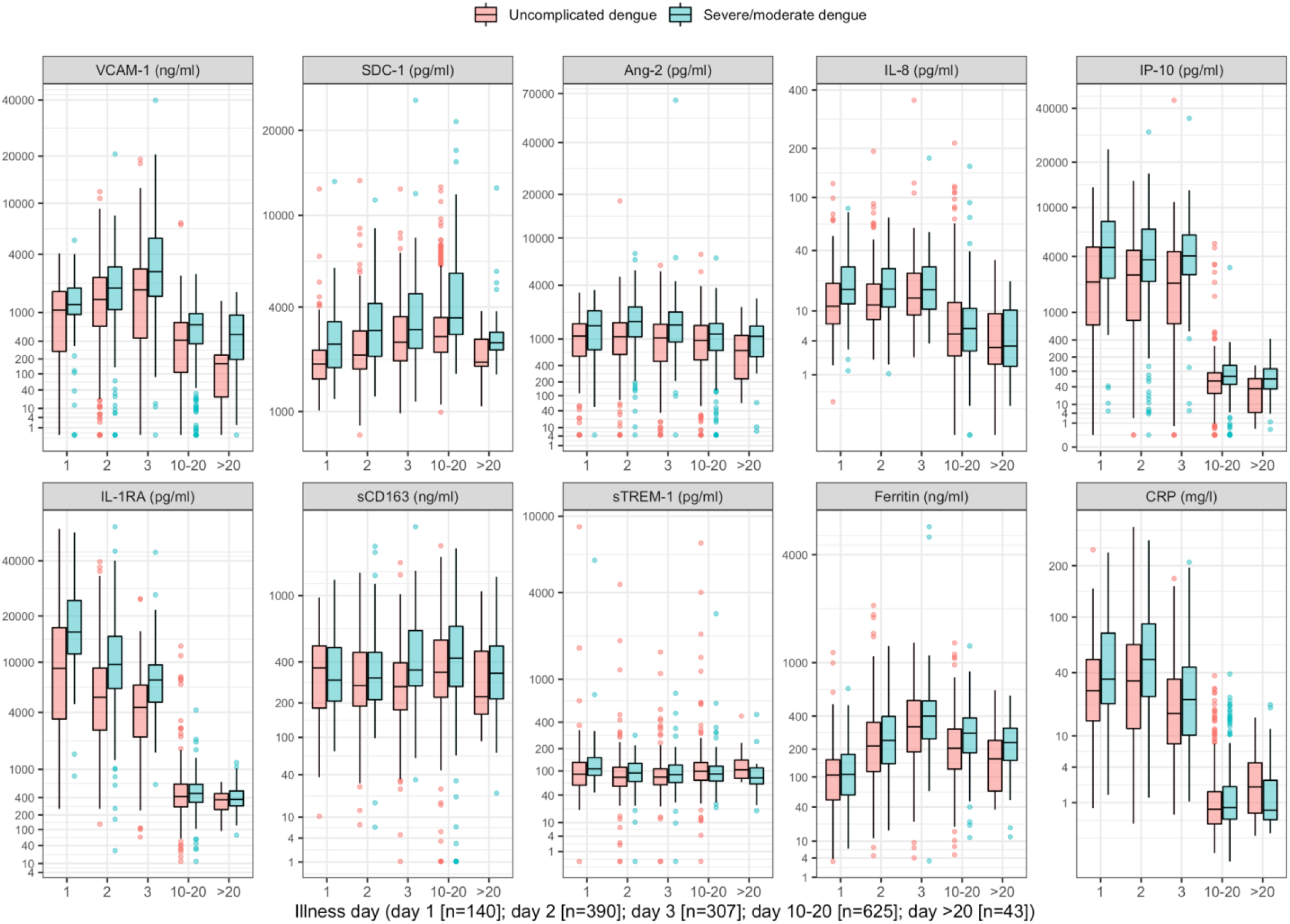
Biomarker levels by groups. VCAM-1: vascular cell adhesion molecule-1; SDC-1: syndecan-1; Ang-2: angiopoietin-2; IL-8: interleukin-8; IP-10: interferon gamma-induced protein-10; IL-1RA: interleukin-1 receptor antagonist; sCD163: soluble cluster of differentiation 163; sTREM-1: soluble triggering receptor expressed on myeloid cells-1; CRP: C-reactive protein Y-axes are transformed using the fourth root transformation

**Table 2.** Summary of biomarkers’ data.

|  | **All patients** |  | **Children** |  | **Adults** |  |
| --- | --- | --- | --- | --- | --- | --- |
|  | **Uncomplicated**<br>**dengue**<br>**(N=556)** | **Severe/moderate**<br>**dengue**<br>**(N=281)** | **Uncomplicated**<br>**dengue**<br>**(N=337)** | **Severe/moderate**<br>**dengue**<br>**(N=127)** | **Uncomplicated**<br>**dengue**<br>**(N=219)** | **Severe/moderate**<br>**dengue**<br>**(N=154)** |
| **At enrollment** |  |  |  |  |  |  |
| VCAM-1 (ng/ml) | 1404 (540, 2548) | 2027 (1122, 3577) | 1442 (447, 2546) | 2020 (1232, 3384) | 1356 (568, 2560) | 2092 (1060, 4202) |
| SDC-1 (pg/ml) | 2334 (1864, 3131) | 2997 (2230, 4201) | 2369 (1861, 3423) | 2846 (2164, 4173) | 2260 (1879, 2898) | 3122 (2278, 4211) |
| Ang-2 (pg/ml) | 1064 (550, 1584) | 1521 (899, 2318) | 1102 (584, 1563) | 1547 (967, 2318) | 944 (516, 1585) | 1516 (885, 2321) |
| IL-8 (pg/ml) | 12 (8, 22) | 17 (11, 28) | 15 (9, 26) | 16 (10, 27) | 10 (7, 15) | 19 (12, 29) |
| IP-10 (pg/ml) | 2502 (732, 4509) | 4092 (2436, 6441) | 2245 (458, 4531) | 3942 (2046, 6287) | 2793 (1370, 4495) | 4242 (2524, 6469) |
| IL-1RA (pg/ml) | 5237 (2603, 9082) | 9105 (5933, 14977) | 4491 (2318, 8977) | 9688 (6109, 16786) | 5721 (3479, 9703) | 8993 (5953, 12935) |
| sCD163 (ng/ml) | 278 (185, 447) | 322 (228, 503) | 326 (212, 481) | 386 (256, 603) | 226 (157, 374) | 291 (207, 410) |
| sTREM-1 (pg/ml) | 81 (59, 114) | 96 (69, 132) | 80 (58, 115) | 93 (67, 128) | 84 (60, 114) | 98 (73, 134) |
| Ferritin (ng/ml) | 233 (116, 406) | 261 (133, 433) | 177 (99, 324) | 224 (110, 402) | 303 (161, 510) | 278 (160, 448) |
| CRP (mg/l) | 25 (10, 54) | 34 (17, 72) | 18 (7, 41) | 24 (13, 58) | 38 (17, 65) | 45 (25, 80) |
| **At follow-up** |  |  |  |  |  |  |
|  | **(N=437)** | **(N=231)** | **(N=292)** | **(N=112)** | **(N=145)** | **(N=119)** |
| VCAM-1 (ng/ml) | 402 (102, 730) | 686 (344, 961) | 579 (182, 858) | 782 (402, 1078) | 173 (26, 388) | 622 (343, 835) |
| SDC-1 (pg/ml) | 2769 (2298, 3514) | 3417 (2815, 5495) | 2957 (2319, 4115) | 3122 (2748, 5507) | 2666 (2196, 3058) | 3745 (2971, 5495) |
| Ang-2 (pg/ml) | 953 (478, 1479) | 1155 (675, 1567) | 1163 (738, 1646) | 1352 (710, 1856) | 565 (302, 923) | 1044 (626, 1345) |
| IL-8 (pg/ml) | 4.9 (2.3, 12.4) | 5.7 (2.7, 10.4) | 6.8 (3.0, 15.1) | 5.9 (2.4, 10.5) | 2.7 (1.6, 4.8) | 5.5 (3.1, 10.4) |
| IP-10 (pg/ml) | 57 (24, 91) | 76 (47, 133) | 67 (33, 98) | 86 (38, 143) | 39 (22, 70) | 75 (48, 108) |
| IL-1RA (pg/ml) | 412 (279, 635) | 455 (328, 626) | 441 (323, 687) | 501 (352, 664) | 336 (210, 480) | 407 (308, 615) |
| sCD163 (ng/ml) | 337 (216, 553) | 412 (257, 661) | 340 (226, 562) | 456 (279, 680) | 328 (199, 523) | 386 (241, 589) |
| sTREM-1 (pg/ml) | 99 (73, 132) | 90 (68, 116) | 98 (72, 132) | 91 (67, 115) | 101 (73, 134) | 90 (70, 116) |
| Ferritin (ng/ml) | 202 (120, 309) | 273 (181, 382) | 177 (112, 263) | 209 (154, 311) | 267 (160, 404) | 322 (247, 436) |
| CRP* (mg/l) | 0.7 (0.3, 1.8) | 0.8 (0.4, 2.0) | 0.6 (0.3, 1.3) | 0.6 (0.3, 1.1) | 1.1 (0.5, 2.7) | 1.1 (0.5, 3.4) |

\*The number of cases with available data for CRP at follow-up in the uncomplicated and severe/moderate dengue groups are 436 and 228 (all patients); 292 and 111 (children); and 218 and 152 (adults) respectively.
VCAM-1: vascular cell adhesion molecule-1; SDC-1: syndecan-1; Ang-2: angiopoietin-2; IL-8: interleukin-8; IP-10: interferon gamma-induced protein-10; IL-1RA: interleukin-1 receptor antagonist; sCD163: soluble cluster of differentiation 163; sTREM-1: soluble triggering receptor expressed on myeloid cells-1; CRP: C-reactive protein
Summary statistics are median (1<sup>st</sup> and 3<sup>rd</sup> quartiles)

### Associations between biomarker levels and the endpoints

In the single models, higher levels of each biomarker on illness day 1, 2 or 3 increased the risk of developing S/MD, with the exception of ferritin in adults where there was a downward trend at higher values (Figure 3, Table 3). We observed differences between children and adults for several biomarkers, the most pronounced being SDC-1, IL-8, ferritin, and IL-1RA. Associations between SDC-1 and IL-8 and the S/MD endpoint were stronger in adults than children, while the effects of IL-1RA and ferritin were stronger in children than adults.

**Figure 3.**
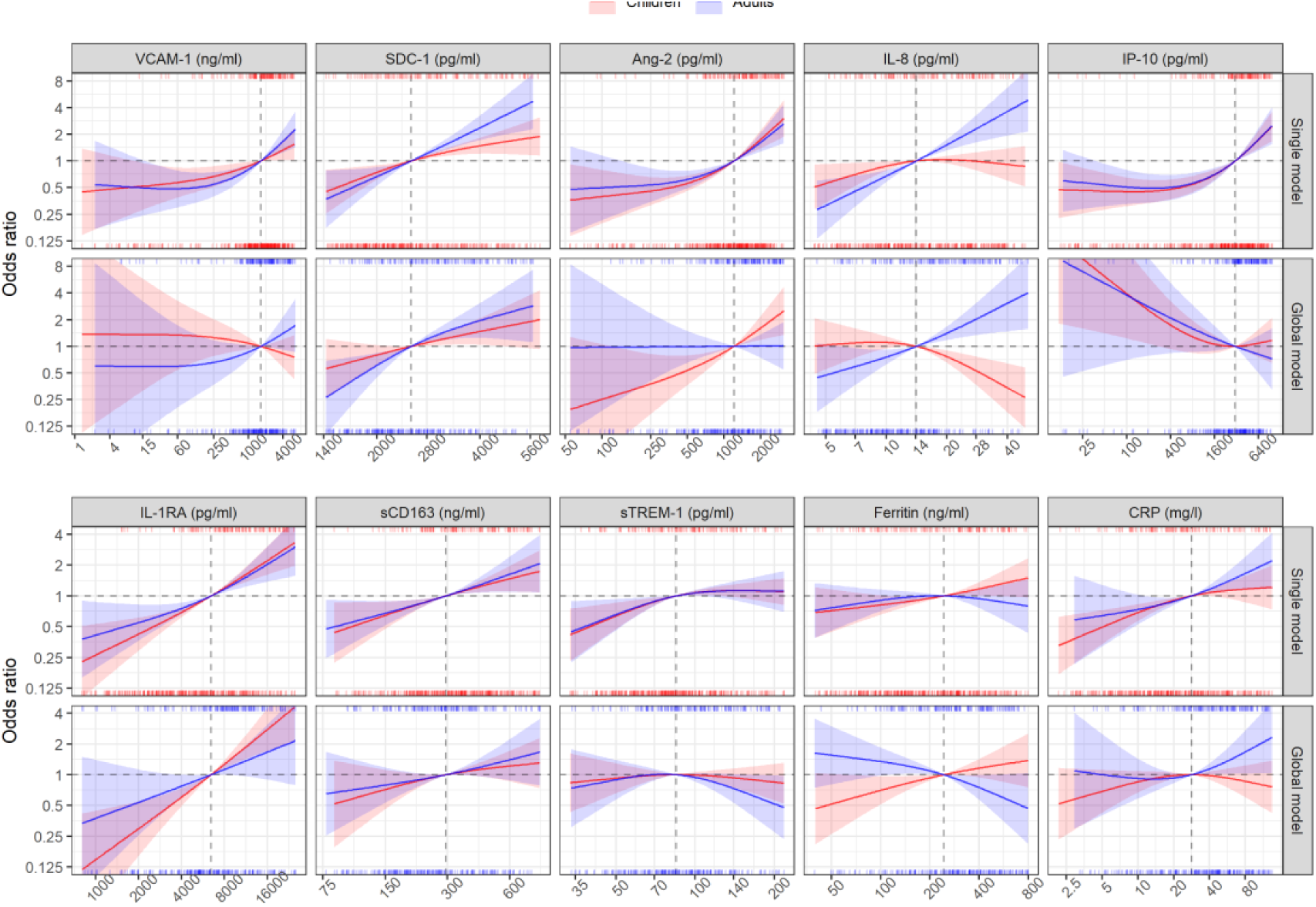
Results from models for the primary endpoint (severe or moderate dengue) The odds ratio of severe/moderate dengue (the red and blue lines) and 95% confidence interval (the red and blue regions) are estimated from multivariable logistic regression models allowing for a non-linear relation of log-2 of the biomarker level with severe/moderate dengue using restricted cubic splines. Each single model contains the corresponding biomarker, age and their interaction, while the global model contains all biomarkers and their interaction with age. The reference values for the odds ratios where the odds ratio is equal to 1, represented by the vertical grey dashed lines, are chosen as the median of the biomarker levels of the whole study population (VCAM-1: 1636 ng/ml; SDC-1: 2519 pg/ml; Ang-2: 1204 pg/ml; IL-8: 14 pg/ml; IP-10: 3093 pg/ml; IL-1RA: 6434 pg/ml; sCD163: 295 ng/ml; sTREM-1: 85 ng/ml; ferritin: 243 ng/ml; and CRP: 28 mg/l). The x-axis represents biomarker levels; it is transformed using log-2 and is truncated by the 5^th^ and 95^th^ percentiles of the biomarker levels of whole study population. The rug plot on the x-axis represents the distribution of individual cases; the bottom rug plot represents the uncomplicated dengue cases and the top rug plot represents the severe/moderate dengue cases (children [<15 years of age] are in red and adults [≥15 years of age] are in blue). The red line and region represent for children, which are estimated at age of 10 years. The blue line and region represent for adults, which are estimated at age of 25 years. VCAM-1: vascular cell adhesion molecule-1; SDC-1: syndecan-1; Ang-2: angiopoietin-2; IL-8: interleukin-8; IP-10: interferon gamma-induced protein-10; IL-1RA: interleukin-1 receptor antagonist; sCD163: soluble cluster of differentiation 163; sTREM-1: soluble triggering receptor expressed on myeloid cells-1; CRP: C-reactive protein

**Table 3.** Results from models for the primary endpoint (severe or moderate dengue)

|  | Single models |  | *P*<sub>overall</sub> | *P*<sub>interaction</sub> | Global model |  | *P*<sub>overall</sub> | *P*<sub>interaction</sub> |
| --- | --- | --- | --- | --- | --- | --- | --- | --- |
|  | Children<br>OR (95% CI) | Adults<br>OR (95% CI) |  |  | Children<br>OR (95% CI) | Adults<br>OR (95% CI) |  |  |
| VCAM-1 (ng/ml) |  |  | <0.001 | 0.715 |  |  | 0.441 | 0.213 |
| - 1636 vs 818 | 1.20 (1.04-1.38) | 1.35 (1.15-1.58) |  |  | 0.90 (0.73-1.10) | 1.22 (0.96-1.57) |  |  |
| - 3272 vs 1636 | 1.25 (1.02-1.53) | 1.48 (1.19-1.85) |  |  | 0.87 (0.66-1.15) | 1.30 (0.93-1.80) |  |  |
| SDC-1 (pg/ml) |  |  | <0.001 | 0.088 |  |  | 0.002 | 0.588 |
| - 2519 vs 1260 | 2.67 (1.31-5.43) | 3.33 (1.32-8.42) |  |  | 2.03 (0.77-5.34) | 5.11 (1.56-16.78) |  |  |
| - 5039 vs 2519 | 1.71 (1.18-2.47) | 3.71 (2.09-6.58) |  |  | 1.76 (0.98-3.14) | 2.52 (1.17-5.42) |  |  |
| Ang-2 (pg/ml) |  |  | <0.001 | 0.524 |  |  | 0.039 | 0.068 |
| - 1204 vs 602 | 1.64 (1.39-1.94) | 1.51 (1.26-1.82) |  |  | 1.67 (1.23-2.25) | 1.01 (0.74-1.38) |  |  |
| - 2409 vs 1204 | 2.21 (1.58-3.10) | 2.00 (1.40-2.85) |  |  | 1.95 (1.25-3.05) | 1.01 (0.65-1.57) |  |  |
| IL-8 (pg/ml) |  |  | <0.001 | <0.001 |  |  | <0.001 | <0.001 |
| - 14 vs 7 | 1.42 (1.05-1.91) | 2.18 (1.47-3.24) |  |  | 0.91 (0.63-1.34) | 1.69 (1.05-2.71) |  |  |
| - 28 vs 14 | 0.99 (0.78-1.25) | 2.33 (1.63-3.33) |  |  | 0.53 (0.36-0.77) | 2.05 (1.34-3.13) |  |  |
| IP-10 (pg/ml) |  |  | <0.001 | 0.984 |  |  | 0.206 | 0.630 |
| - 3093 vs 1546 | 1.46 (1.26-1.68) | 1.45 (1.21-1.73) |  |  | 0.94 (0.73-1.19) | 0.80 (0.57-1.12) |  |  |
| - 6186 vs 3093 | 1.68 (1.35-2.09) | 1.69 (1.29-2.22) |  |  | 1.08 (0.77-1.51) | 0.82 (0.52-1.29) |  |  |
| IL-1RA (pg/ml) |  |  | <0.001 | 0.082 |  |  | <0.001 | 0.032 |
| - 6434 vs 3217 | 1.69 (1.42-2.03) | 1.48 (1.21-1.81) |  |  | 2.07 (1.52-2.84) | 1.45 (0.98-2.15) |  |  |
| - 12868 vs 6434 | 1.82 (1.46-2.27) | 1.70 (1.29-2.24) |  |  | 2.16 (1.53-3.05) | 1.47 (0.94-2.30) |  |  |
| sCD163 (ng/ml) |  |  | <0.001 | 0.551 |  |  | 0.217 | 0.341 |
| - 295 vs 147 | 1.57 (1.14-2.15) | 1.49 (1.13-1.98) |  |  | 1.40 (0.89-2.22) | 1.27 (0.84-1.91) |  |  |
| - 589 vs 295 | 1.46 (1.10-1.93) | 1.61 (1.09-2.37) |  |  | 1.21 (0.87-1.69) | 1.39 (0.89-2.18) |  |  |
| sTREM-1 (pg/ml) |  |  | 0.059 | 0.997 |  |  | 0.555 | 0.393 |
| - 85 vs 42 | 1.87 (1.23-2.84) | 1.79 (1.10-2.93) |  |  | 1.13 (0.70-1.81) | 1.21 (0.65-2.26) |  |  |
| - 169 vs 85 | 1.12 (0.91-1.38) | 1.12 (0.82-1.53) |  |  | 0.89 (0.65-1.21) | 0.61 (0.38-0.99) |  |  |
| Ferritin (ng/ml) |  |  | 0.042 | 0.054 |  |  | 0.008 | 0.002 |
| - 243 vs 122 | 1.18 (1.01-1.38) | 1.06 (0.89-1.27) |  |  | 1.30 (1.04-1.64) | 0.78 (0.61-0.99) |  |  |
| - 487 vs 243 | 1.26 (1.00-1.58) | 0.90 (0.66-1.23) |  |  | 1.22 (0.89-1.67) | 0.66 (0.44-1.00) |  |  |
| CRP (mg/l) |  |  | <0.001 | 0.031 |  |  | 0.184 | 0.138 |
| - 28 vs 14 | 1.26 (1.12-1.41) | 1.25 (1.03-1.52) |  |  | 1.08 (0.93-1.25) | 1.10 (0.85-1.44) |  |  |
| - 56 vs 28 | 1.13 (0.95-1.34) | 1.38 (1.11-1.71) |  |  | 0.93 (0.75-1.15) | 1.36 (1.02-1.81) |  |  |

2 *P*<sub>overall</sub> is derived from Wald test for the overall association of the biomarker with the endpoint; *P*<sub>interaction</sub> is from the test for the interaction between the biomarker and age.
3 The odds ratios are estimated at age of 10 and 25 years, represented as children and adults respectively.

In the global model there were some differences compared to the single models (Figure 3, Table 3). The biomarkers SDC-1 and IL-1RA were the most stable relative to the single models for both children and adults. However, for IP-10 the trend of the association with S/MD changed from positive to negative in both children and adults. In children, VCAM-1 changed the trend from positive to weakly negative and IL-8 changed the trend from weakly positive to negative. Other biomarkers showed weaker associations with the endpoint in the global model based on the ORs. In addition, the differences of the associations between children and adults were more marked, particularly for Ang-2, IL-8 and ferritin.

Similar patterns were observed in the various analyses related to the secondary endpoints, as described in detail in the appendix 5.

### Best combinations of biomarkers associated with the primary endpoint

For children, the best subset that showed the clearest association with S/MD was the combination of the six markers IL-1RA, Ang-2, IL-8, ferritin, IP-10, and SDC-1 with an AIC of 465.9. This model was selected most often in the bootstrap procedure, but was not highly robust (it was selected in 134 of the 1000 samples) (Tables 4, S5). Over the 1000 samples, the six variables had an inclusion frequency ranging from 73.5% for SDC-1 to 100% for IL-1RA (Table S6). The best combination of two biomarkers was IL-1RA and ferritin, the best of three added Ang-2, the best of four added IP-10, and the best of five added IL-8. The best combinations of two and five variables were most robust with a selection percentage of 43.7% and 44%. The best of five had almost the same AIC as the best subset of six markers (467.6 versus 465.9) (Table 4). The coefficients of the selected biomarkers were similar to the initial model estimates (Table S6).

**Table 4.** Best combinations of biomarkers associated with severe or moderate dengue for children.

|  | Best of all combinations | Best combination of 2 variables | Best combination of 3 variables | Best combination of 4 variables | Best combination of 5 variables |
| --- | --- | --- | --- | --- | --- |
| Variables |  |  |  |  |  |
| - VCAM-1 |  |  |  |  |  |
| - SDC-1 | + |  |  |  |  |
| - Ang-2 | + |  | + | + | + |
| - IL-8 | + |  |  |  | + |
| - IP-10 | + |  |  | + | + |
| - IL-1RA | + | + | + | + | + |
| - sCD163 |  |  |  |  |  |
| - sTREM-1 |  |  |  |  |  |
| - Ferritin | + | + | + | + | + |
| - CRP |  |  |  |  |  |
| AIC of the selected model | 465.9 | 484.7 | 480.0 | 473.7 | 467.6 |
| Bootstrap results |  |  |  |  |  |
| - Model selection frequency, *n* (%) | 134 (13.4) | 437 (43.7) | 239 (23.9) | 317 (31.7) | 440 (44.0) |
| - Rank by selection frequency of the selected model | 1 | 1 | 2 | 1 | 1 |

*VCAM-1: vascular cell adhesion molecule-1; SDC-1: syndecan-1; Ang-2: angiopoietin-2; IL-8: interleukin-8; IP-10: interferon gamma-induced protein-10; IL-1RA: interleukin-1* *receptor antagonist; sCD163: soluble cluster of differentiation 163; sTREM-1: soluble triggering receptor expressed on myeloid cells-1; CRP: C-reactive protein; AIC: Akaike* *information criterion*

For adults, the best subset included the seven markers SDC-1, IL-8, ferritin, sTREM-1, IL-1RA, IP-10, and sCD163. This model was selected 79 times among 1000 bootstrap samples, but still was selected more often than the other models (Tables 5, S7). Over the 1000 samples, the seven variables had a bootstrap inclusion frequency ranging from 59.1% for sCD163 to 99.2% for SDC-1 (Table S8). The best combination of two biomarkers included SDC-1 and IL-8, the best of three added ferritin, the best of four added IL-1RA, and the best of five added sTREM-1. The best combination of two was the most robust with a selection percentage of 56.7%, followed by the best of three variables (43.2%) (Table 5). The coefficients of the selected markers were also similar to the initial model estimates (Table S8).

**Table 5.**
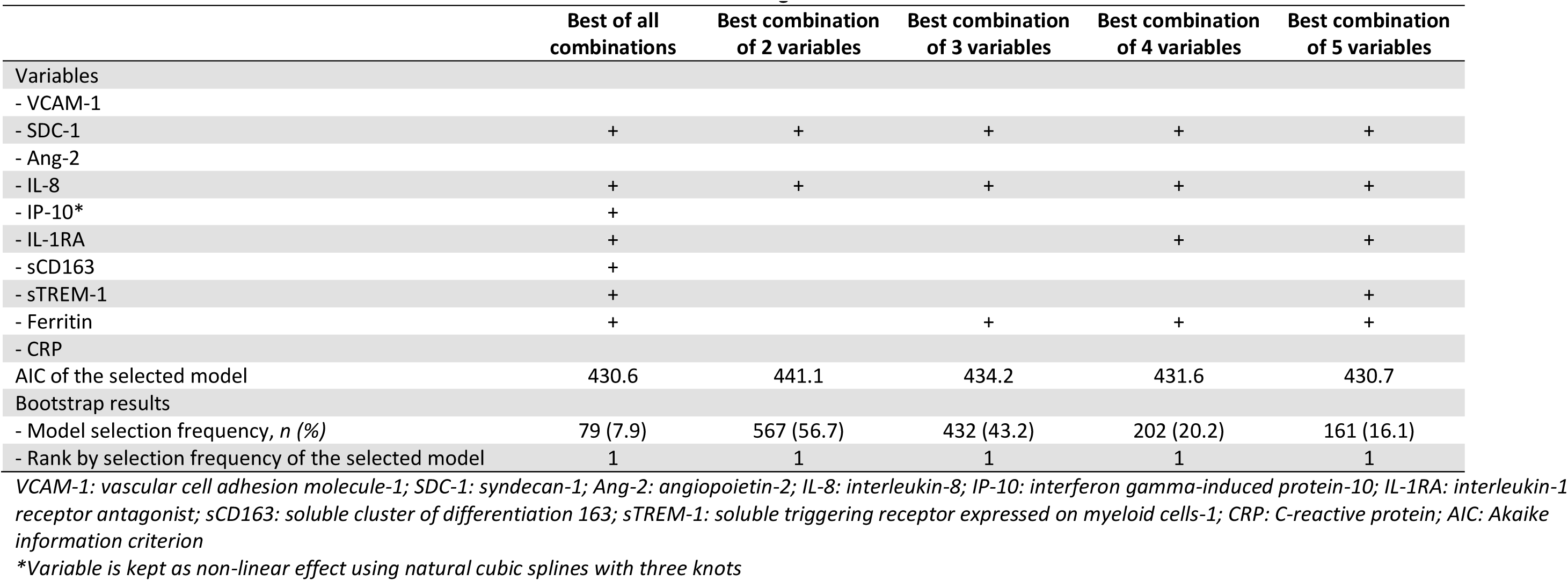
Best combinations of biomarkers associated with severe or moderate dengue for adults.

|  | Best of all combinations | Best combination of 2 variables | Best combination of 3 variables | Best combination of 4 variables | Best combination of 5 variables |
| --- | --- | --- | --- | --- | --- |
| Variables |  |  |  |  |  |
| - VCAM-1 |  |  |  |  |  |
| - SDC-1 | + | + | + | + | + |
| - Ang-2 |  |  |  |  |  |
| - IL-8 | + | + | + | + | + |
| - IP-10* | + |  |  |  |  |
| - IL-1RA | + |  |  | + | + |
| - sCD163 | + |  |  |  |  |
| - sTREM-1 | + |  |  |  | + |
| - Ferritin | + |  | + | + | + |
| - CRP |  |  |  |  |  |
| AIC of the selected model | 430.6 | 441.1 | 434.2 | 431.6 | 430.7 |
| Bootstrap results |  |  |  |  |  |
| - Model selection frequency, *n* (%) | 79 (7.9) | 567 (56.7) | 432 (43.2) | 202 (20.2) | 161 (16.1) |
| - Rank by selection frequency of the selected model | 1 | 1 | 1 | 1 | 1 |

*VCAM-1: vascular cell adhesion molecule-1; SDC-1: syndecan-1; Ang-2: angiopoietin-2; IL-8: interleukin-8; IP-10: interferon gamma-induced protein-10; IL-1RA: interleukin-1* *receptor antagonist; sCD163: soluble cluster of differentiation 163; sTREM-1: soluble triggering receptor expressed on myeloid cells-1; CRP: C-reactive protein; AIC: Akaike* *information criterion*
*\*Variable is kept as non-linear effect using natural cubic splines with three knots*

## Discussion

This nested case-control study has shown that a range of endothelial, immune activation and inflammatory biomarkers measured during the early febrile phase of dengue are associated with progression to worse clinical outcomes in both children and adults. In children we found IL-1RA to have the most robust association with S/MD, whereas in adults we found SDC-1 and IL-8 to have the most robust association. For children, the best combination (ordered by robustness) included six biomarkers IL-1RA, Ang-2, IL-8, ferritin, IP-10, and SDC-1; for adults the best combination identified comprised seven biomarkers SDC-1, IL-8, ferritin, sTREM-1, IL-1RA, IP-10, and sCD163.

These results are important, as they not only provide novel information on the utility of certain biomarkers alone and in combination for prediction of dengue progression using data from a large multi-country cohort study across endemic settings, but also demonstrate some key differences between paediatric and adult dengue. Clinical phenotypes of dengue in children and adults differ, with children experiencing more shock and adults more organ impairment and bleeding, with distinct clinical management guidelines published by the WHO. Our results imply dengue pathogenesis may differ by age, with distinct combinations of immune-activation and vascular markers demonstrated between children and adults. Hence patients’ age should be considered when developing biomarker panels for dengue risk prediction. Our results are directly applicable to the development of point-of-care panels capable of multiplex analysis and suited for use in outpatient settings for dengue prognosis, with scope for incorporation with innovative point-of-care technologies. Balancing model fit, robustness, and parsimony, we suggest the combination of five biomarkers IL-1RA, Ang-2, IL-8, ferritin, and IP-10 for children, and the combination of three biomarkers SDC-1, IL-8, and ferritin for adults to be used in practice.

Our results add to the current literature on biomarkers in severe/moderate dengue compared with uncomplicated dengue, by including early time-points prior to the development of the severe manifestations as well as providing data on the use of biomarker combinations, which takes into consideration the complex immune-inflammatory-vascular pathogenesis of severe dengue. We observed that there were marked changes in the associations between individual biomarkers and outcomes when considering them together, while other biomarkers showed consistent associations. For example, the associations of SDC-1 and IL-1RA with S/MD were mostly similar in the single and global models, which suggests that they might be mediators in the pathway of the other biomarkers that we considered to severe outcomes.

The use of biomarker panels for the prediction of severe outcomes in dengue has been investigated in previous studies, using several statistical approaches.[40–44] However, because of small sample size and differences in the biomarkers assessed, the associations found vary between studies and as yet there are no validated prognostic panels for dengue. Dengue cases are forecasted to increase over the next few decades and, given the limited healthcare resources available in many endemic settings, particularly during epidemics, there is an urgent need to develop innovative methods to rapidly identify patients likely to develop complications and require hospital care.[45] Previously we showed that CRP as a single biomarker was useful for early dengue diagnosis and risk prediction, which is currently easy to use in all settings.[35] However, future point-of-care testing could be improved by using a combination biomarkers outlined in this study. With the advent of novel technologies including microarray platforms and multiplex lateral flow assays, the cost is likely to come down in the future, allowing for wide-spread use in low-to-middle-income countries.

Methods of variable selection have been discussed previously but there remains no clear consensus regarding the best approach.[46,47] We adopted a data-driven ‘best subset’ approach which we think offers advantages over other methods, given the complexity of the biomarkers involved and their interactions. We also explored other approaches for variable selection,[46–48] and the results were very similar. In children, four methods (backward elimination, forward selection, stepwise forward, and stepwise backward) resulted in the same six selected biomarkers as the best subset method. Augmented backward elimination added another biomarker (VCAM-1), while Bayesian projection eliminated one (SDC-1) (Table S9). In adults, two methods (backward elimination and stepwise backward) gave the same results as the best subset method. Forward selection and stepwise forward identified the same biomarkers as the best subset of five biomarkers. Augmented backward elimination also added VCAM-1, while Bayesian projection selected only two biomarkers, which were the same as the best subset of two (Table S10).

Strengths of our study include the large sample size and use of a nested case-control dataset from a prospective multi-country cohort study with consistent data collection and standardized outcome definitions and laboratory methodologies. The biomarker panel we selected was guided by pathogenesis studies, focusing on pathways activated early in the disease course, thus ensuring clinical relevance.

There are some limitations in our study. One being we analysed the biomarkers at only one time-point in the early phase; limited financial resources did not allow us to evaluate the full range of biomarkers across the whole IDAMS population and at more time-points. Secondly, we did not assess differences by serotype and primary/secondary infection due to a limited sample size.

In conclusion, higher levels of the ten biomarkers (VCAM-1, SDC-1, Ang-2, IL-8, IP-10, IL-1RA, sCD163, sTREM-1, ferritin, and CRP), when considered individually, are associated with increased risk of adverse clinical outcomes in both children and adults with dengue. The best biomarker combination for children includes IL-1RA, Ang-2, IL-8, ferritin, IP-10, and SDC-1; for adults, SDC-1, IL-8, ferritin, sTREM-1, IL-1RA, IP-10, and sCD163 were selected. These findings serve to assist the development of biomarker panels to improve future triage and early assessment of dengue patients. This would aid not only individual patient management and facilitate healthcare allocation which would be of major public health benefit especially in outbreak settings, but could also serve as potential biological endpoints for dengue clinical trials.

## Data Availability

All data referred to in the manuscript are available in the link below.

https://doi.org/10.5287/bodleian:JN2wXDpjq

## List of abbreviations

AIC: Akaike information criterion
Ang-2: angiopoietin-2
CRP: C-reactive protein
DENV: dengue virus
ELISA: enzyme-linked immunosorbent assay
IL-1RA: interleukin-1 receptor antagonist
IL-8: interleukin-8
IP-10: interferon gamma-induced protein-10
OR: odds ratio
OUCRU: Oxford University Clinical Research Unit
RT-PCR: reverse transcriptase polymerase chain reaction
S/MD: severe and moderate dengue
sCD163: soluble cluster of differentiation 163
SDC-1: syndecan-1
sTREM-1: soluble triggering receptor expressed on myeloid cells-1
VCAM-1: vascular cell adhesion molecule-1
WHO: World Health Organization

## Acknowledgements

We would like to acknowledge all the patients who took part in this study and the medical and nursing staff who looked helped in their management, at all the participating hospitals in Vietnam, Cambodia, Malaysia, and El Salvador.

## Appendix 1. Laboratory evaluation of the ten biomarkers

All research blood samples enrolled at different study sites were processed at site laboratories within one hour after drawn from participants with the same procedure. The blood samples were centrifuged at 500 g/min in 10 min at 4°C, collected plasma, and stored at −20°C. The specimens enrolled from international sites were transported on dry ice to the Oxford University Clinical Research Unit (OUCRU) laboratory by worldwide couriers. Biomarker levels were measured on these stored samples at two time-points: enrollment sample (illness day 1-3) and follow-up (day 10-31 post symptom onset) using the quantitative magnetic bead assays and enzyme-linked immunosorbent assays (ELISAs).

Eight biomarkers (VCAM-1, SDC-1, Ang-2, IL-8, IP-10, IL-1RA, sCD163 and sTREM-1) combined in a premixed magnetic bead panel (Cat No. LXSAHM; R&D) was selected to investigate these interested targets. A commercial kit used for CRP level measurements was Human Cardiovascular Disease (CVD) Magnetic Bead Panel 3 (Cat. No. HCVD3MAG-67K) produced by EMD Millipore Corporation. Each assay had two quality controls (low and high levels) which was acquired along with the standards and unknown specimens.

The xPonent 3.1 software installed in the Luminex200^TM^ analyzer was used to acquire and analyze the data of the magnetic luminex assays. The system was calibrated daily using the Luminex calibration (Cat. No. LX200-CAL-K25) and verification kits (Cat. No. LX200-CON-K25). Equipment settings included probe height adjustment, number of events for each analyte, sample size, gate settings, and bead set were followed as the kit’s instruction. A background was set up using assay buffer instead of sample in all luminex assays.

As recommended by manufacturers, the magnetic bead assays were performed using samples with less than two freeze/thaw cycles and the samples after thawed completely were centrifuged to remove particles prior to use in the assays. The samples were diluted at the different dilutions in order to fall within the standard curve range in each assay.

The magnetic bead assays were designed in multiplex sandwich ELISAs. The magnetic microparticles pre-coated with specific antibodies were pipetted into the wells containing standards or controls or diluted samples and the immobilized antibodies bound the analytes of interest for two hours incubation at room temperature or overnight (16-18 hours) at 4°C on a shaker. Then, these analytes were detected specifically by a secondary biotinylated antibody cocktail during the next incubation. After being washed to remove any unbound antibody, the streptavidin-phycoerythrin conjugate was added to bind to the biotinylated antibody. Finally, the microparticles were re-suspended in buffer and read using the Luminex200 analyzer. The microparticles were re-suspended immediately prior to reading by shaking the plate for two minutes on the shaker.

Ferritin levels were measured separately using Human Ferritin ELISA kit (Cat. No. ARG80501, Arigo). This assay is a quantitative sandwich ELISA. An antibody specific for Ferritin was pre-coated onto a microplate plate. Standard or samples were pipetted into the wells and any Ferritin present was bound by the immobilized antibody. After washing away any unbound substances, a horseradish peroxidase (HRP) conjugated antibody specific for ferritin was added to each well and incubated. A substrate solution (3,3′,5,5′-tetramethylbenzidine [TMB]) was then added to the wells and color developed in proportion to the amount of ferritin bound in the initial step. The color development was stopped by addition of acid and the intensity of the color was measured by a wavelength of 450 nm. The concentration of ferritin in the sample was then determined by comparing the optical density of samples to the standard curve.

## Appendix 2. Clinical endpoint definition

**Table S1.** Definition of severe and moderate dengue components.

| Endpoint | Definition |
| --- | --- |
| Severe plasma leakage | Dengue shock syndrome or respiratory distress due to plasma leakage |
| Moderate plasma leakage | Did not fulfill criteria for severe plasma leakage and had at least one of the following criteria: (1) maximum haematocrit change was 20% or more, and (2) having evidence of fluid accumulation |
| Severe bleeding | Any bleeding into a critical organ or required any blood transfusion of packed red cells or whole blood without pre-anaemia, or bleeding with complication |
| Moderate bleeding | Did not fulfill criteria for severe bleeding and had at least one of the following criteria: (1) severe bleeding by clinical judgement, (2) bleeding required any blood transfusion other than packed red cells or whole blood, (3) bleeding required other intervention (e.g. nasal packing, cross-match, etc.), and (4) receiving packed red cells or whole blood with pre-existing anaemia and with a consistent haemoglobin value |
| Severe neurologic involvement | Abnormal neurologic examination and neurologic involvement that resulted in death or ongoing sequelae that impaired daily function, or required intubation, shunting or intensive care |
| Moderate neurology involvement | Single convulsion without hospitalization or other complication |
| Severe hepatic involvement | Jaundice or coagulopathy or encephalopathy |
| Moderate hepatic involvement | Any alanine aminotransferase (ALT) or aspartate aminotransferase (AST) result of 400 IU/L or more |
| Severe other major organ failure | Creatine kinase or other enzymes (e.g. troponin) abnormalities and functional abnormalities (e.g. reduced cardiac ejection fraction less than 50% or new electrocardiogram [ECG] abnormalities) or required specific intervention (e.g. inotropic support) |
| Moderate other major organ failure | Troponin abnormalities alone or creatine kinase abnormalities without cardiac ejection fraction less than 50% |

## Appendix 3. Statistical analysis

### Treatment of values lower than the limit of detection

There were several biomarker values lower than the limit of detection (LOD): 9% for VCAM-1, 5% for Ang-2, 1% for IP-10 and sTREM-1, <1% for IL-8 and sCD163, and none for SDC-1, IL-1RA, ferritin, and CRP. All of them were set at the LOD and a dummy binary variable (Yes/No) was created for each biomarker to describe whether the value was lower than the LOD or not. In all models for the first aim (to investigate the association of biomarkers with clinical outcomes), for each biomarker that had values below a LOD, we included the binary variable “<LOD” as a covariate.

### Analysis of the secondary endpoints: case-control setting

Since cases and controls were selected based on the primary endpoint, in the analyses of the secondary endpoints we lose the case-control distinction. We used inverse probability weighting (IPW) to correct for different inclusion probabilities between controls and cases in our data set:[1,2] the weight of all cases was 1, while the weight of the controls was the inverse of the inclusion probability in each country (Vietnam: 1301/436; Cambodia: 272/39; Malaysia: 230/58; and El Salvador: 288/23). A robust (sandwich) estimate of the standard error was used for estimating 95% confidence intervals. For the severe dengue (SD) endpoint, the non-linear effect was not considered because of the low number of events.

### Analysis to find the best combination of biomarkers to predict the primary endpoint (aim #2)

The results from the ‘single models’ and ‘global model’ in the first aim showed that the association between the biomarkers and the primary endpoint differed by age. We therefore performed the analysis separately for children (<15 years of age) and adults (≥15 years of age). The procedure was done in two steps. In step #1 we built an ‘initial model’ including all biomarkers, but possibly with less flexible structure than the global model. In step #2 we determined the best combination of biomarkers from the initial models defined in step #1.

**Step #1:** As the number of primary endpoint events was limited (127 in children and 154 in adults), we tried to keep the events-per-variable (EPV) at more than 10 by including only important terms (all the ten biomarkers but only some of the non-linear trends and binary variables that represent values <LOD).[3] For each biomarker, we fitted and compared four logistic regression models:

1. model with the biomarker with a linear effect as the only covariate: logit(Y) = α + β*X
2. model with the biomarker with a non-linear effect using restricted cubic splines (as in the single and global model): logit(Y) = α + β_1_*spline(X)_1_ + β_2_*spline(X)_2_
3. model with the biomarker with a linear effect and the dummy binary variable for value <LOD: logit(Y) = α + β*X + γ*(X<LOD)
4. model with the biomarker with a non-linear effect and the dummy binary variable for value <LOD: logit(Y) = α + β_1_*spline(X)_1_ + β_2_*spline(X)_2_ + γ*(X<LOD)

### Y is the primary endpoint, X is a biomarker

We calculated and compared the Akaike information criterion (AIC) of these four models and included the non-linear effect and/or additional binary variable of values <LOD only if it had the lowest AIC and this value was at least 5 lower than for model (1). The AICs of these models are summarized in the table below:

| Biomarker | Children |  |  |  | Adults |  |  |  |
| --- | --- | --- | --- | --- | --- | --- | --- | --- |
|  | model1 | model2 | model3 | model4 | model1 | model2 | model3 | model4 |
| VCAM-1 | **530.8** | 531.3 | 530.6 | 532.6 | **499.8** | 496.7 | 501.3 | 496.4 |
| SDC-1 | **537.9** | 538.0 | - | - | **459.5** | 461.0 | - | - |
| Ang-2 | **511.0** | 509.6 | 512.5 | 509.7 | **493.5** | 490.8 | 493.3 | 492.8 |
| IL-8 | **548.5** | 549.0 | 548.1 | 546.8 | **457.7** | 457.8 | 456.5 | 457.8 |
| IP-10 | **521.2** | 517.2 | 523.0 | 518.3 | 500.7 | **492.6** | 502.5 | 494.5 |
| IL-1RA | **492.9** | 494.9 | - | - | **493.5** | 494.2 | - | - |
| sCD163 | **531.9** | 533.2 | 531.9 | 533.2 | **497.6** | 499.3 | 499.5 | 501.1 |
| sTREM-1 | **545.7** | 544.2 | 545.6 | 545.4 | **507.8** | 509.5 | 507.0 | 505.0 |
| Ferritin | **542.6** | 544.4 | - | - | **509.3** | 505.2 | - | - |
| CRP | **536.7** | 536.4 | - | - | **505.1** | 506.8 | - | - |

*The selected models are in bold face*

The initial model for children included all biomarkers as a linear term without any binary variable for values <LOD, and the final initial model for adults included IP-10 with a non-linear term and all the other biomarkers with a linear term, again without any binary variable for values <LOD. The number of parameters of the initial model was 10 and 11 for children and adults; the EPV was then 12.7 and 14 respectively.

**Step #2:** From the ‘initial model’, we performed several approaches to find the best combination of biomarkers associated with the primary endpoint.

The primary method was the ‘best subset’ approach, in which all possible subsets of biomarkers (2^10^ = 1024 subsets) were evaluated and compared via the AIC. The subset with the lowest AIC was selected as the best subset, we also determined the best subset of exactly 2, 3, 4, and 5 biomarkers.

We also performed other approaches to investigate whether they gave similar results. These included backward elimination, forward selection, stepwise forward, stepwise backward, augmented backward elimination, and Bayesian projection variable selection.[3–5]

### Checking model robustness by bootstrap resampling

To check the robustness (stability) of the selected ‘best subset’ model, we used a bootstrap procedure by resampling with replacement from the original data set (1000 times). For each bootstrap sample, we performed the ‘best subset’ approach (similar to step #2 above) to determine the best model based on the lowest AIC. From the 1000 samples we calculated:[3]

i. Inclusion frequency for each of the ten biomarkers
ii. The root mean squared difference (RMSD) ratio of each regression coefficient. The root mean squared difference is computed between the 1000 estimates of the regression coefficient after the best subset selection and its value in the initial model (which includes all 10 biomarkers and is estimated on the original data).

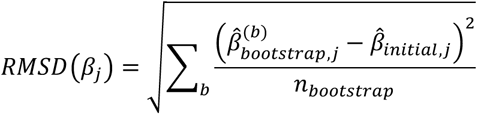

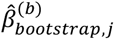 is the estimate of parameter j in bootstrap sample b
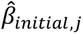 is the estimate of parameter j based on the initial model
*n_bootstrap_* is the number of bootstrap samples (1000) The RMSD ratio is the RMSD divided by the standard error of that coefficient in the initial model.
iii. Relative bias conditional on selection for each parameter

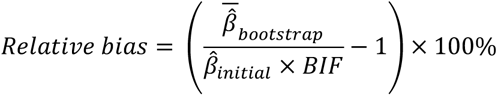

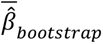 is the mean bootstrapped estimate of the parameter
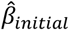 is the initial model estimate of the parameter
*BIF* is the bootstrap inclusion frequency of the corresponding biomarker
iv. The selection frequencies for the finally selected model and the 20 most frequent selected models.
v. The median and the 2.5^th^ and 97.5^th^ percentiles of the regression coefficient of each biomarker.

In ii, iii and v, the value of a parameter was set at zero if the marker was not selected

## Appendix 4. Additional descriptive analysis

**Figure S1.**
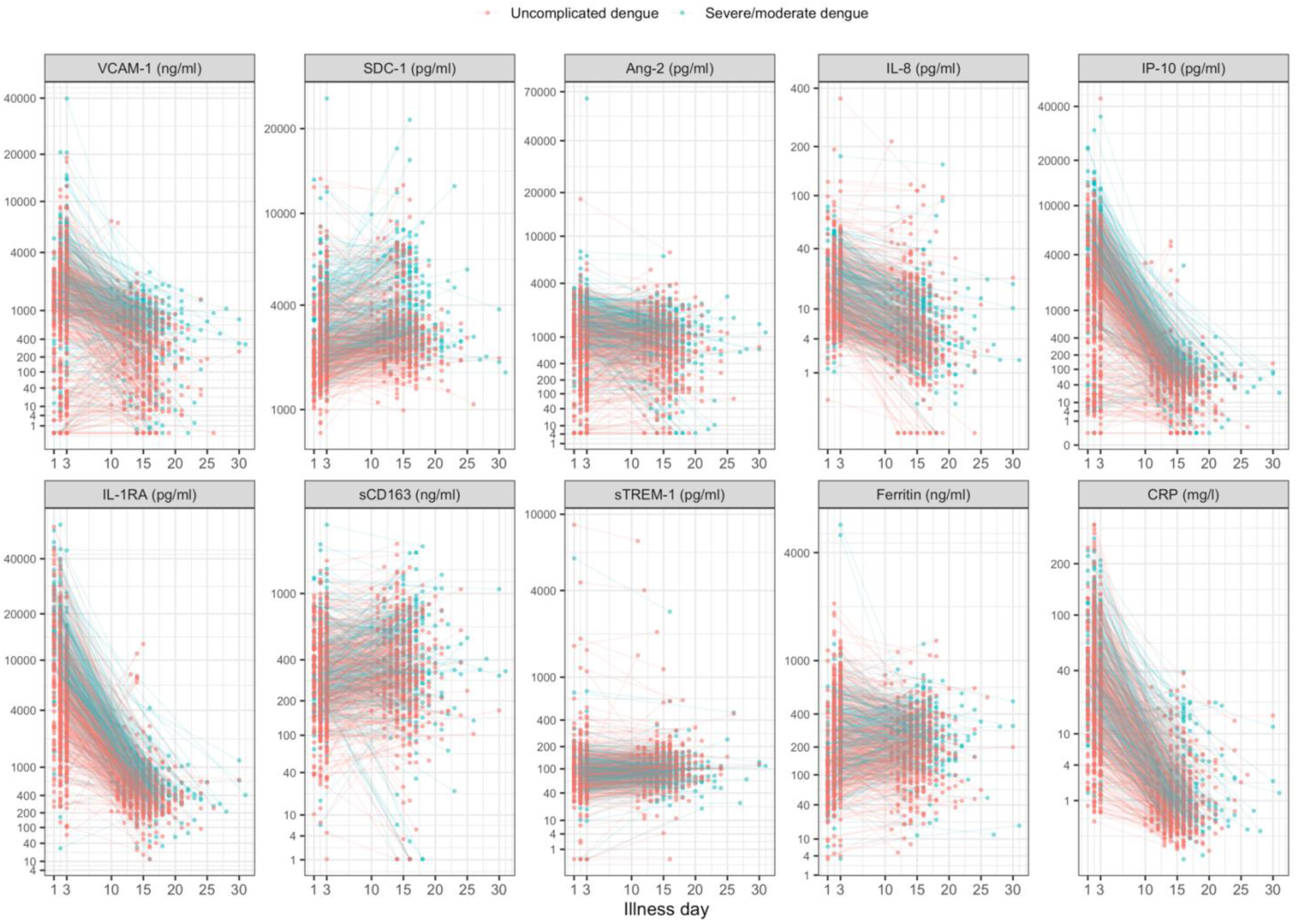
Biomarker levels by individual. Y-axes are transformed using the fourth root transformation

**Figure S2.**
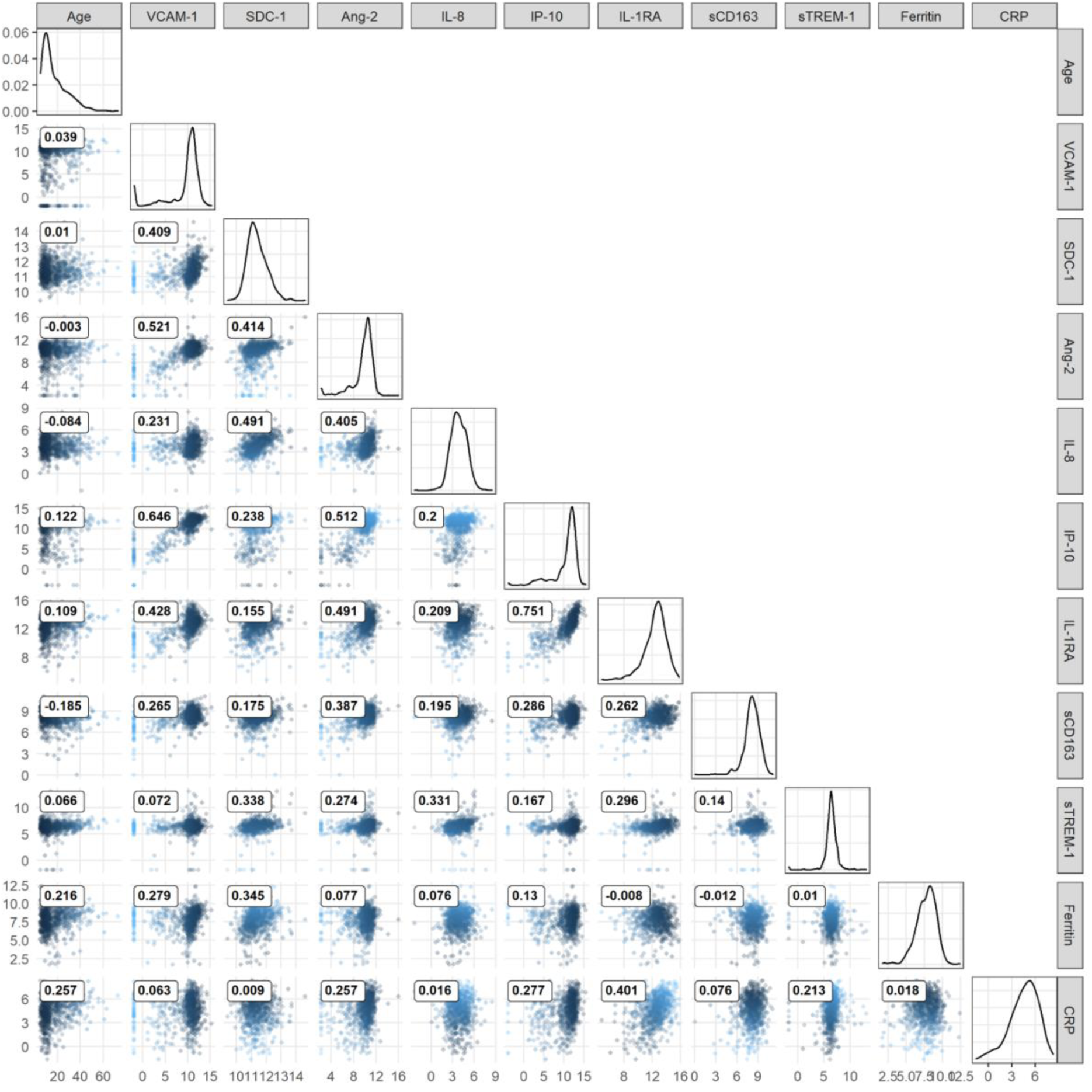
Pairwise correlation of biomarker levels at enrollment and age. All biomarker levels were transformed using log-2. The number inside each scatter plot represents the Spearman’s rank correlation coefficient of the two variables at the corresponding column and row. When the column and row refer to the same variable, the corresponding scatter plot is replaced by a density plot to reflect the distribution of that biomarker.

## Appendix 5. Results for secondary endpoints

In the single models, higher levels of the biomarkers generally increased the risk of developing severe dengue (SD), however, as the number of events was small, the confidence intervals (CIs) were wide and the association was not certain (Figure S3, Table S2). For severe dengue or dengue with warning signs (SD/DWWS) and hospitalization endpoints, the associations were similar to the primary endpoint, apart for sCD163, sTREM-1, and CRP (Figures S4-S5, Tables S3-S4) – these biomarkers did not show an association with endpoints. Moreover, the odds ratios (ORs) of SD/DWWS and hospitalization were generally lower than of severe or moderate dengue (S/MD) for every 2-fold difference in biomarker levels.

The difference between the global and single models in the analysis of secondary endpoints was similar to in the primary endpoint. The most stable biomarkers were SDC-1 and IL-1RA, while IP-10 markedly changed the trend of the association with the endpoints; others showed a weaker association.

**Figure S3.**
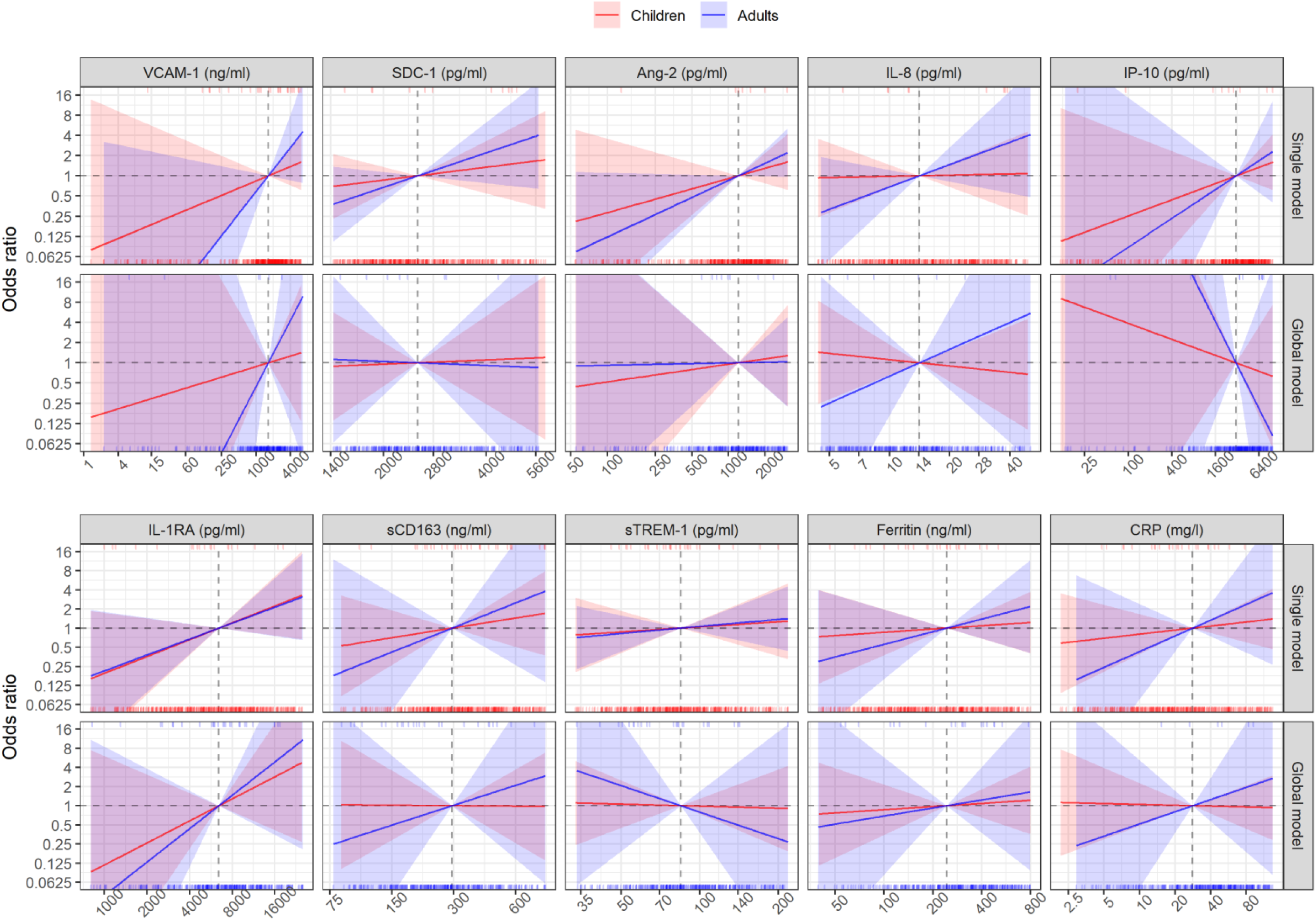
Results from models for severe dengue endpoint.

**Table S2.** Results from models for severe dengue endpoint.

|  | Single models |  |  |  | Global model |  |  |  |
| --- | --- | --- | --- | --- | --- | --- | --- | --- |
|  | Children<br>OR (95% CI) | Adults<br>OR (95% CI) | *P*<sub>overall</sub> | *P*<sub>interaction</sub> | Children<br>OR (95% CI) | Adults<br>OR (95% CI) | *P*<sub>overall</sub> | *P*<sub>interaction</sub> |
| VCAM-1 (ng/ml) | 1.28 (0.77-2.12) | 2.13 (0.88-5.15) | 0.236 | 0.202 | 1.20 (0.35-4.12) | 3.13 (0.19-50.77) | 0.723 | 0.491 |
| SDC-1 (pg/ml) | 1.55 (0.41-5.96) | 3.26 (0.69-15.52) | 0.307 | 0.438 | 1.16 (0.12-11.07) | 0.87 (0.03-27.93) | 0.987 | 0.884 |
| Ang-2 (pg/ml) | 1.42 (0.70-2.88) | 1.79 (0.97-3.31) | 0.148 | 0.580 | 1.20 (0.33-4.31) | 1.02 (0.33-3.19) | 0.961 | 0.830 |
| IL-8 (pg/ml) | 1.04 (0.47-2.33) | 2.15 (0.67-6.88) | 0.420 | 0.338 | 0.81 (0.28-2.31) | 2.51 (0.16-38.43) | 0.741 | 0.446 |
| IP-10 (pg/ml) | 1.32 (0.75-2.33) | 1.64 (0.58-4.62) | 0.443 | 0.708 | 0.76 (0.17-3.30) | 0.22 (0.01-7.12) | 0.693 | 0.473 |
| IL-1RA (pg/ml) | 1.84 (0.81-4.16) | 1.78 (0.80-3.94) | 0.098 | 0.956 | 2.22 (0.51-9.64) | 3.35 (0.45-24.87) | 0.386 | 0.696 |
| sCD163 (ng/ml) | 1.43 (0.52-3.94) | 2.43 (0.27-21.70) | 0.617 | 0.645 | 0.98 (0.27-3.59) | 2.06 (0.15-27.47) | 0.855 | 0.590 |
| sTREM-1 (pg/ml) | 1.20 (0.44-3.28) | 1.29 (0.55-3.05) | 0.793 | 0.914 | 0.93 (0.30-2.88) | 0.38 (0.02-8.76) | 0.827 | 0.541 |
| Ferritin (ng/ml) | 1.13 (0.59-2.16) | 1.58 (0.59-4.23) | 0.659 | 0.501 | 1.12 (0.55-2.29) | 1.34 (0.25-7.02) | 0.926 | 0.819 |
| CRP (mg/l) | 1.16 (0.71-1.88) | 1.77 (0.56-5.62) | 0.620 | 0.418 | 0.97 (0.58-1.63) | 1.55 (0.33-7.32) | 0.773 | 0.500 |

*Odds ratios (95% confidence intervals) are calculated for each 2-fold increase of the biomarkers and are estimated at age of 10 and 25 years, represented as children and adults respectively; *P*<sub>overall</sub> is derived from Wald test for the overall association of the biomarker with the endpoint; *P*<sub>interaction</sub> is from the test for the interaction between the biomarker and age.*

**Figure S4.**
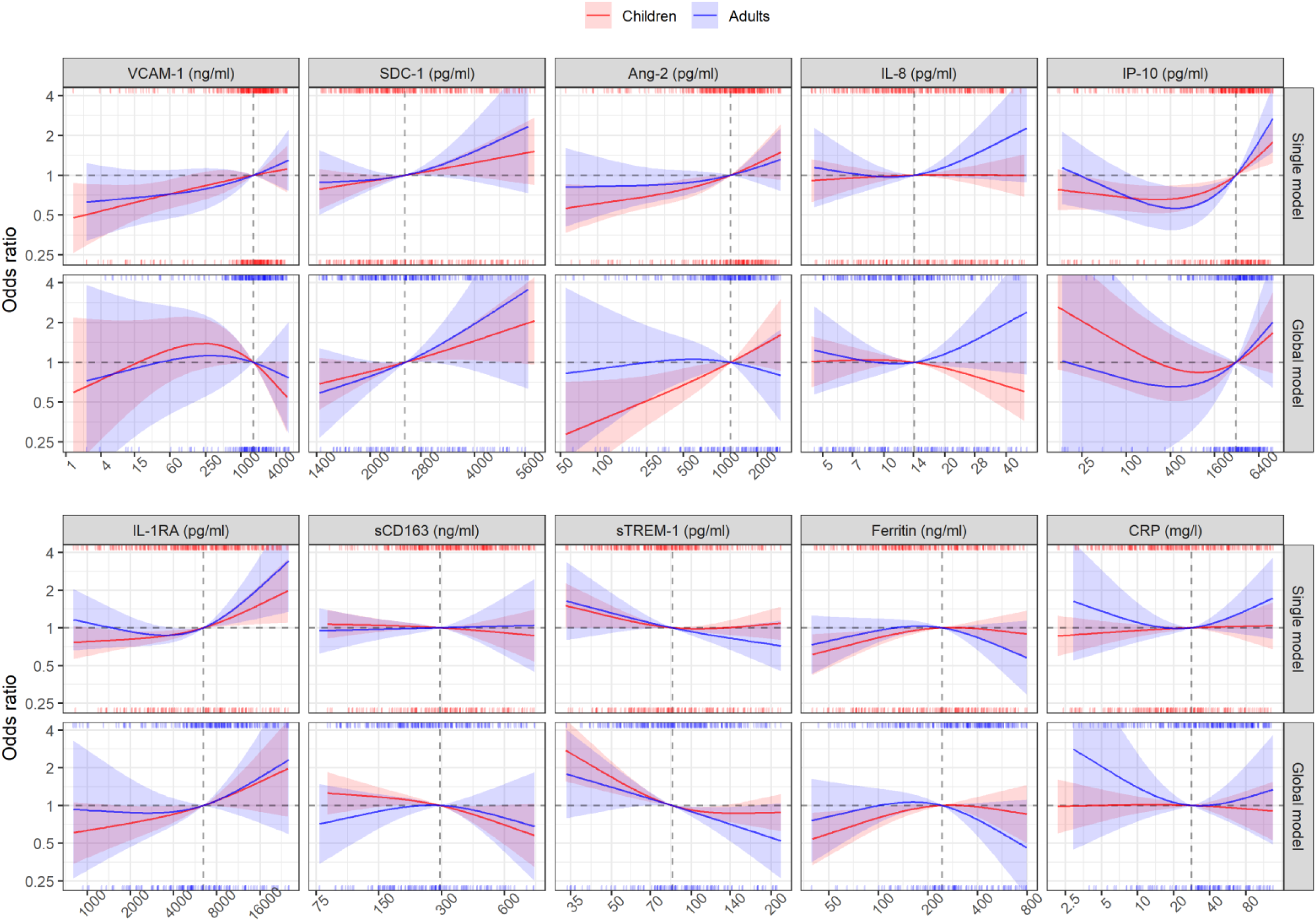
Results from models for severe dengue or dengue with warning signs endpoint.

**Table S3.** Results from models for severe dengue or dengue with warning signs endpoint.

|  | Single models |  |  |  | Global model |  |  |  |
| --- | --- | --- | --- | --- | --- | --- | --- | --- |
| | Children<br>OR (95% CI) | Adults<br>OR (95% CI) | $P_{overall}$ | $P_{interaction}$ | Children<br>OR (95% CI) | Adults<br>OR (95% CI) | $P_{overall}$ | $P_{interaction}$ |
| VCAM-1 (ng/ml) |  |  | 0.025 | 0.374 |  |  | 0.469 | 0.763 |
| - 1636 vs 818 | 1.06 (0.93-1.22) | 1.11 (0.92-1.33) |  |  | 0.82 (0.66-1.02) | 0.92 (0.66-1.30) |  |  |
| - 3272 vs 1636 | 1.06 (0.87-1.29) | 1.13 (0.88-1.46) |  |  | 0.74 (0.55-1.01) | 0.88 (0.56-1.40) |  |  |
| SDC-1 (pg/ml) |  |  | 0.032 | 0.363 |  |  | 0.116 | 0.773 |
| - 2519 vs 1260 | 1.34 (0.86-2.09) | 1.15 (0.56-2.33) |  |  | 1.57 (0.89-2.75) | 1.89 (0.71-5.03) |  |  |
| - 5039 vs 2519 | 1.40 (0.90-2.17) | 2.00 (0.79-5.08) |  |  | 1.78 (1.00-3.18) | 2.88 (0.71-11.69) |  |  |
| Ang-2 (pg/ml) |  |  | 0.008 | 0.637 |  |  | 0.009 | 0.011 |
| - 1204 vs 602 | 1.23 (1.05-1.44) | 1.12 (0.93-1.36) |  |  | 1.37 (1.07-1.75) | 0.95 (0.69-1.30) |  |  |
| - 2409 vs 1204 | 1.34 (0.95-1.88) | 1.22 (0.82-1.80) |  |  | 1.42 (0.91-2.21) | 0.85 (0.48-1.51) |  |  |
| IL-8 (pg/ml) |  |  | 0.040 | 0.020 |  |  | 0.053 | 0.030 |
| - 14 vs 7 | 1.05 (0.87-1.27) | 0.97 (0.67-1.41) |  |  | 0.96 (0.77-1.21) | 0.94 (0.62-1.42) |  |  |
| - 28 vs 14 | 1.01 (0.85-1.19) | 1.45 (0.94-2.22) |  |  | 0.78 (0.62-0.99) | 1.48 (0.89-2.44) |  |  |
| IP-10 (pg/ml) |  |  | <0.001 | 0.176 |  |  | 0.059 | 0.537 |
| - 3093 vs 1546 | 1.26 (1.09-1.44) | 1.44 (1.16-1.80) |  |  | 1.16 (0.87-1.55) | 1.30 (0.81-2.09) |  |  |
| - 6186 vs 3093 | 1.39 (1.13-1.71) | 1.75 (1.26-2.44) |  |  | 1.33 (0.89-1.99) | 1.49 (0.78-2.87) |  |  |
| IL-1RA (pg/ml) |  |  | 0.005 | 0.381 |  |  | 0.425 | 0.955 |
| - 6434 vs 3217 | 1.17 (1.06-1.30) | 1.15 (0.97-1.36) |  |  | 1.24 (1.01-1.52) | 1.14 (0.80-1.63) |  |  |
| - 12868 vs 6434 | 1.37 (1.06-1.77) | 1.70 (1.13-2.55) |  |  | 1.38 (0.93-2.05) | 1.45 (0.80-2.64) |  |  |
| sCD163 (ng/ml) |  |  | 0.854 | 0.719 |  |  | 0.193 | 0.419 |
| - 295 vs 147 | 0.96 (0.84-1.08) | 1.03 (0.85-1.25) |  |  | 0.85 (0.71-1.03) | 1.12 (0.81-1.55) |  |  |
| - 589 vs 295 | 0.91 (0.69-1.22) | 1.03 (0.61-1.73) |  |  | 0.71 (0.50-1.00) | 0.80 (0.44-1.47) |  |  |
| sTREM-1 (pg/ml) |  |  | 0.221 | 0.472 |  |  | 0.002 | 0.132 |
| - 85 vs 42 | 0.75 (0.56-1.00) | 0.69 (0.41-1.16) |  |  | 0.48 (0.32-0.73) | 0.64 (0.36-1.16) |  |  |
| - 169 vs 85 | 1.04 (0.84-1.28) | 0.78 (0.57-1.07) |  |  | 0.87 (0.69-1.10) | 0.62 (0.39-1.00) |  |  |
| Ferritin (ng/ml) |  |  | 0.034 | 0.258 |  |  | 0.024 | 0.075 |
| - 243 vs 122 | 1.13 (1.02-1.26) | 1.01 (0.85-1.19) |  |  | 1.17 (1.01-1.35) | 0.96 (0.76-1.23) |  |  |
| - 487 vs 243 | 0.96 (0.77-1.20) | 0.76 (0.53-1.08) |  |  | 0.94 (0.71-1.25) | 0.67 (0.42-1.07) |  |  |
| CRP (mg/l) |  |  | 0.747 | 0.622 |  |  | 0.662 | 0.448 |
| - 28 vs 14 | 1.03 (0.96-1.12) | 0.97 (0.79-1.20) |  |  | 0.99 (0.89-1.10) | 0.84 (0.59-1.20) |  |  |
| - 56 vs 28 | 1.02 (0.87-1.19) | 1.19 (0.92-1.54) |  |  | 0.96 (0.79-1.17) | 1.06 (0.75-1.49) |  |  |

Odds ratios are estimated at age of 10 and 25 years, represented as children and adults respectively; $P_{overall}$ is derived from Wald test for the overall association of the biomarker with the endpoint; $P_{interaction}$ is from the test for the interaction between the biomarker and age

**Figure S5.**
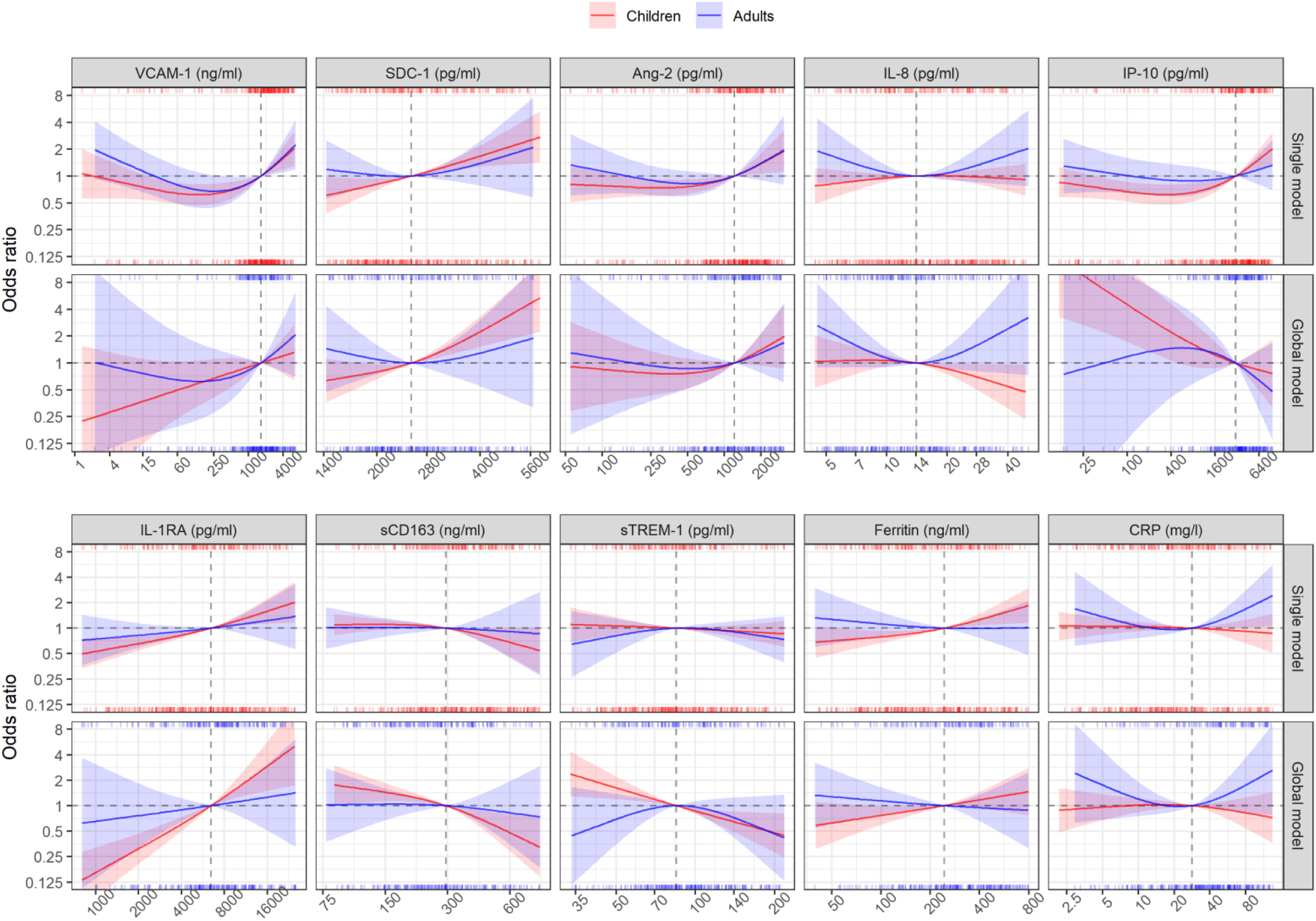
Results from models for hospitalization endpoint.

**Table S4.** Results from models for hospitalization endpoint.

|  | Single models |  | *P*<sub>overall</sub> | *P*<sub>interaction</sub> | Global model |  | *P*<sub>overall</sub> | *P*<sub>interaction</sub> |
| --- | --- | --- | --- | --- | --- | --- | --- | --- |
|  | Children<br>OR (95% CI) | Adults<br>OR (95% CI) |  |  | Children<br>OR (95% CI) | Adults<br>OR (95% CI) |  |  |
| VCAM-1 (ng/ml) |  |  | <0.001 | 0.009 |  |  | 0.092 | 0.137 |
| - 1636 vs 818 | 1.28 (1.11-1.49) | 1.28 (1.03-1.60) |  |  | 1.16 (0.90-1.48) | 1.28 (0.87-1.90) |  |  |
| - 3272 vs 1636 | 1.42 (1.14-1.76) | 1.46 (1.08-1.97) |  |  | 1.16 (0.81-1.64) | 1.41 (0.84-2.37) |  |  |
| SDC-1 (pg/ml) |  |  | <0.001 | 0.187 |  |  | 0.006 | 0.406 |
| - 2519 vs 1260 | 1.82 (1.01-3.28) | 0.79 (0.31-2.05) |  |  | 1.70 (0.84-3.41) | 0.62 (0.16-2.49) |  |  |
| - 5039 vs 2519 | 2.22 (1.36-3.63) | 1.81 (0.65-5.07) |  |  | 3.70 (1.90-7.22) | 1.66 (0.39-7.07) |  |  |
| Ang-2 (pg/ml) |  |  | 0.012 | 0.337 |  |  | 0.497 | 0.789 |
| - 1204 vs 602 | 1.27 (1.07-1.52) | 1.21 (0.91-1.62) |  |  | 1.27 (0.92-1.77) | 1.16 (0.76-1.77) |  |  |
| - 2409 vs 1204 | 1.58 (1.08-2.32) | 1.61 (0.86-3.04) |  |  | 1.63 (0.90-2.93) | 1.45 (0.70-3.01) |  |  |
| IL-8 (pg/ml) |  |  | 0.007 | 0.002 |  |  | 0.024 | 0.021 |
| - 14 vs 7 | 1.14 (0.90-1.44) | 0.73 (0.47-1.15) |  |  | 0.94 (0.66-1.33) | 0.63 (0.36-1.13) |  |  |
| - 28 vs 14 | 0.98 (0.81-1.18) | 1.32 (0.85-2.05) |  |  | 0.70 (0.50-0.97) | 1.59 (0.81-3.12) |  |  |
| IP-10 (pg/ml) |  |  | 0.002 | 0.242 |  |  | 0.005 | 0.212 |
| - 3093 vs 1546 | 1.32 (1.13-1.54) | 1.10 (0.86-1.40) |  |  | 0.80 (0.56-1.14) | 0.77 (0.45-1.30) |  |  |
| - 6186 vs 3093 | 1.50 (1.18-1.90) | 1.17 (0.81-1.70) |  |  | 0.84 (0.51-1.40) | 0.66 (0.32-1.35) |  |  |
| IL-1RA (pg/ml) |  |  | <0.001 | 0.685 |  |  | <0.001 | 0.389 |
| - 6434 vs 3217 | 1.31 (1.17-1.46) | 1.13 (0.94-1.36) |  |  | 2.05 (1.57-2.66) | 1.18 (0.73-1.89) |  |  |
| - 12868 vs 6434 | 1.41 (1.11-1.80) | 1.17 (0.79-1.72) |  |  | 2.25 (1.39-3.64) | 1.19 (0.62-2.28) |  |  |
| sCD163 (ng/ml) |  |  | 0.208 | 0.722 |  |  | 0.007 | 0.419 |
| - 295 vs 147 | 0.90 (0.79-1.04) | 0.98 (0.76-1.27) |  |  | 0.69 (0.53-0.89) | 0.96 (0.62-1.50) |  |  |
| - 589 vs 295 | 0.69 (0.47-1.00) | 0.91 (0.46-1.82) |  |  | 0.49 (0.30-0.80) | 0.83 (0.36-1.91) |  |  |
| sTREM-1 (pg/ml) |  |  | 0.635 | 0.371 |  |  | 0.011 | 0.053 |
| - 85 vs 42 | 0.93 (0.67-1.29) | 1.35 (0.71-2.58) |  |  | 0.53 (0.34-0.81) | 1.74 (0.67-4.52) |  |  |
| - 169 vs 85 | 0.89 (0.71-1.13) | 0.83 (0.54-1.28) |  |  | 0.55 (0.36-0.83) | 0.57 (0.26-1.29) |  |  |
| Ferritin (ng/ml) |  |  | <0.001 | 0.011 |  |  | 0.129 | 0.117 |
| - 243 vs 122 | 1.22 (1.08-1.38) | 0.92 (0.72-1.18) |  |  | 1.23 (1.02-1.49) | 0.91 (0.68-1.21) |  |  |
| - 487 vs 243 | 1.40 (1.10-1.79) | 0.99 (0.68-1.46) |  |  | 1.25 (0.89-1.75) | 0.93 (0.54-1.59) |  |  |
| CRP (mg/l) |  |  | 0.379 | 0.190 |  |  | 0.139 | 0.053 |
| - 28 vs 14 | 0.97 (0.89-1.06) | 1.01 (0.83-1.24) |  |  | 0.97 (0.86-1.11) | 0.95 (0.71-1.27) |  |  |
| - 56 vs 28 | 0.95 (0.78-1.15) | 1.35 (1.01-1.81) |  |  | 0.89 (0.69-1.15) | 1.37 (0.88-2.13) |  |  |

Odds ratios are estimated at age of 10 and 25 years, represented as children and adults respectively; *P*<sub>overall</sub> is derived from Wald test for the overall association of the biomarker with the endpoint; *P*<sub>interaction</sub> is from the test for the interaction between the biomarker and age

## Appendix 6. Results from bootstrap resampling to check model robustness

Tables S5-S8 show the robustness (stability) of the selected models. The best subset models for children and adults based on the original data ranked first (Tables S5 and S7), but they were selected in only 13.4% and 7.9% of the resamples, indicating the instability of these models. The almost equal inclusion frequencies of the models ranked next suggest that there are many competing models of the selected one. Variable selection also added to uncertainty about the regression coefficients of the parameters, which is evidenced by the RMSD ratio of more than 1 in most of the biomarkers, except for sTREM-1 (0.92) and CRP (0.88) in children (Tables S6 and S8). Regarding relative conditional bias, which quantifies expected bias induced by variable selection of a parameter when it is selected, it is quite small for the first 3-4 selected parameters (IL-1RA, Ang-2, IL-8, and ferritin for children, and SDC-1, IL-8, and ferritin for adults), all of which have bootstrap inclusion percentages greater than 90%. This bias is much higher in the parameters for which selection is less certain. The bootstrap median and percentiles of the regression coefficients of the biomarkers reflect the variability of the coefficients over the different models selected in the bootstrap samples. The coefficients of the selected parameters from the initial estimates and the bootstrap median were very similar, suggesting no selection bias in the selected model.

**Table S5.** Model selection frequencies for children.

| Model | Included variables |  |  |  |  |  |  |  |  |  | Count | Percent |
| --- | --- | --- | --- | --- | --- | --- | --- | --- | --- | --- | --- | --- |
|  | VCAM-1 | SDC-1 | Ang-2 | IL-8 | IP-10 | IL-1RA | sCD163 | sTREM-1 | Ferritin | CRP |  |  |
| **1** |  | + | + | + | + | + |  |  | + |  | **134** | **13.4** |
| 2 |  | + | + | + | + | + | + |  | + |  | 100 | 10.0 |
| 3 |  |  | + | + | + | + | + |  | + |  | 55 | 5.5 |
| 4 |  |  | + | + | + | + |  |  | + |  | 54 | 5.4 |
| 5 | + | + | + | + |  | + | + |  | + |  | 48 | 4.8 |
| 6 | + | + | + | + | + | + |  |  | + |  | 47 | 4.7 |
| 7 | + | + | + | + | + | + | + |  | + |  | 46 | 4.6 |
| 8 | + | + | + | + |  | + |  |  | + |  | 40 | 4.0 |
| 9 |  | + | + | + | + | + |  |  | + | + | 39 | 3.9 |
| 10 |  | + | + | + | + | + | + | + | + |  | 36 | 3.6 |
| 11 |  | + | + | + | + | + | + |  | + | + | 28 | 2.8 |
| 12 |  | + | + | + | + | + |  | + | + |  | 23 | 2.3 |
| 13 | + |  | + | + | + | + |  |  | + |  | 23 | 2.3 |
| 14 | + |  | + | + |  | + |  |  | + |  | 17 | 1.7 |
| 15 |  | + | + | + | + | + |  | + | + | + | 15 | 1.5 |
| 16 | + | + | + | + |  | + |  |  | + | + | 14 | 1.4 |
| 17 |  |  | + | + | + | + |  |  | + | + | 13 | 1.3 |
| 18 | + |  | + | + | + | + | + |  | + |  | 12 | 1.2 |
| 19 | + | + | + | + | + | + |  | + | + |  | 12 | 1.2 |
| 20 |  |  | + | + | + | + | + |  | + | + | 11 | 1.1 |

*Selected model is ranked first (bold face)*

**Table S6.**
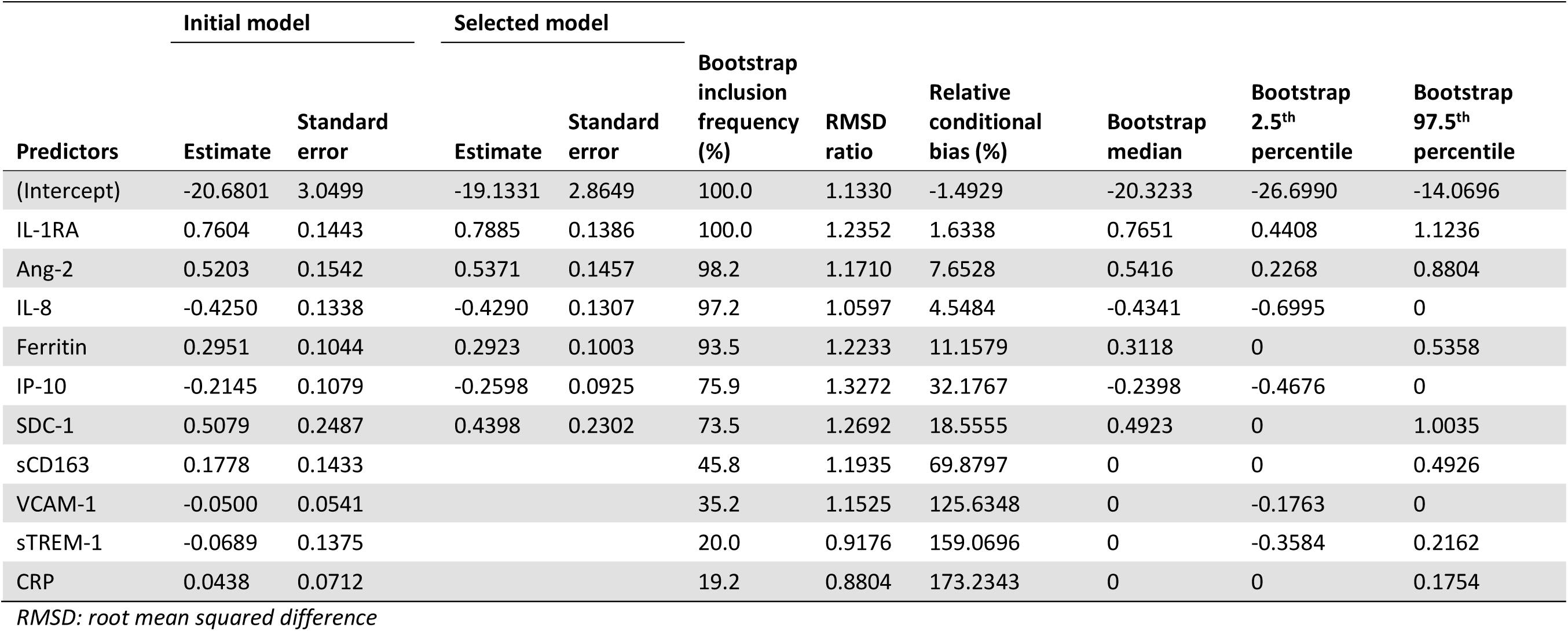
Model stability for children.

| Predictors | Initial model |  | Selected model |  | Bootstrap inclusion frequency (%) | RMSD ratio | Relative conditional bias (%) | Bootstrap median | Bootstrap 2.5<sup>th</sup> percentile | Bootstrap 97.5<sup>th</sup> percentile |
| --- | --- | --- | --- | --- | --- | --- | --- | --- | --- | --- |
|  | Estimate | Standard error | Estimate | Standard error |  |  |  |  |  |  |
| (Intercept) | -20.6801 | 3.0499 | -19.1331 | 2.8649 | 100.0 | 1.1330 | -1.4929 | -20.3233 | -26.6990 | -14.0696 |
| IL-1RA | 0.7604 | 0.1443 | 0.7885 | 0.1386 | 100.0 | 1.2352 | 1.6338 | 0.7651 | 0.4408 | 1.1236 |
| Ang-2 | 0.5203 | 0.1542 | 0.5371 | 0.1457 | 98.2 | 1.1710 | 7.6528 | 0.5416 | 0.2268 | 0.8804 |
| IL-8 | -0.4250 | 0.1338 | -0.4290 | 0.1307 | 97.2 | 1.0597 | 4.5484 | -0.4341 | -0.6995 | 0 |
| Ferritin | 0.2951 | 0.1044 | 0.2923 | 0.1003 | 93.5 | 1.2233 | 11.1579 | 0.3118 | 0 | 0.5358 |
| IP-10 | -0.2145 | 0.1079 | -0.2598 | 0.0925 | 75.9 | 1.3272 | 32.1767 | -0.2398 | -0.4676 | 0 |
| SDC-1 | 0.5079 | 0.2487 | 0.4398 | 0.2302 | 73.5 | 1.2692 | 18.5555 | 0.4923 | 0 | 1.0035 |
| sCD163 | 0.1778 | 0.1433 |  |  | 45.8 | 1.1935 | 69.8797 | 0 | 0 | 0.4926 |
| VCAM-1 | -0.0500 | 0.0541 |  |  | 35.2 | 1.1525 | 125.6348 | 0 | -0.1763 | 0 |
| sTREM-1 | -0.0689 | 0.1375 |  |  | 20.0 | 0.9176 | 159.0696 | 0 | -0.3584 | 0.2162 |
| CRP | 0.0438 | 0.0712 |  |  | 19.2 | 0.8804 | 173.2343 | 0 | 0 | 0.1754 |

*RMSD: root mean squared difference*

**Table S7.**
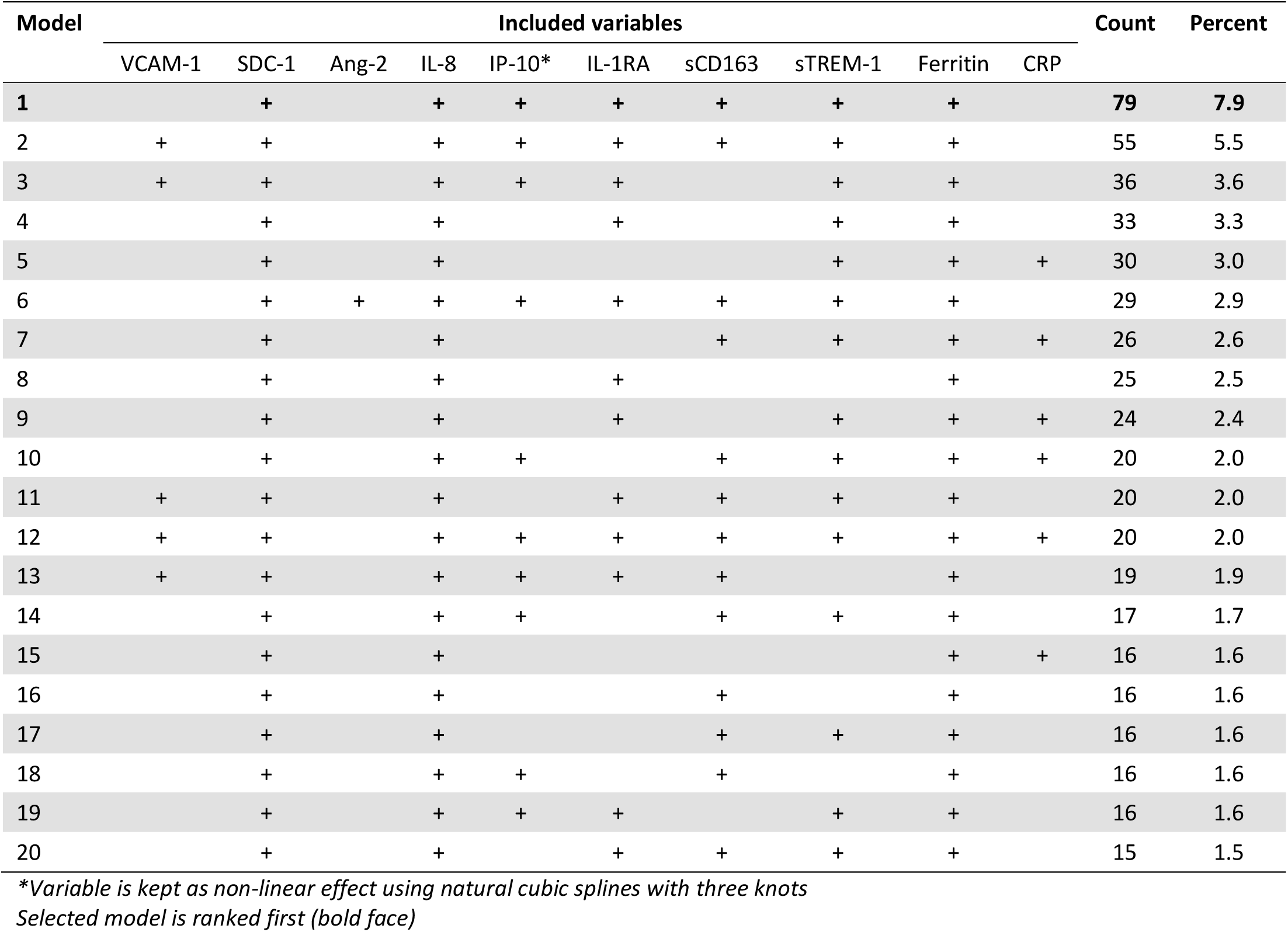
Model selection frequencies for adults.

| Model | Included variables |  |  |  |  |  |  |  |  |  | Count | Percent |
| --- | --- | --- | --- | --- | --- | --- | --- | --- | --- | --- | --- | --- |
|  | VCAM-1 | SDC-1 | Ang-2 | IL-8 | IP-10* | IL-1RA | sCD163 | sTREM-1 | Ferritin | CRP |  |  |
| **1** |  | + |  | + | + | + | + | + | + |  | **79** | **7.9** |
| 2 | + | + |  | + | + | + | + | + | + |  | 55 | 5.5 |
| 3 | + | + |  | + | + | + |  | + | + |  | 36 | 3.6 |
| 4 |  | + |  | + |  | + |  | + | + |  | 33 | 3.3 |
| 5 |  | + |  | + |  |  |  | + | + | + | 30 | 3.0 |
| 6 |  | + | + | + | + | + | + | + | + |  | 29 | 2.9 |
| 7 |  | + |  | + |  |  | + | + | + | + | 26 | 2.6 |
| 8 |  | + |  | + |  | + |  |  | + |  | 25 | 2.5 |
| 9 |  | + |  | + |  | + |  | + | + | + | 24 | 2.4 |
| 10 |  | + |  | + | + |  | + | + | + | + | 20 | 2.0 |
| 11 | + | + |  | + |  | + | + | + | + |  | 20 | 2.0 |
| 12 | + | + |  | + | + | + | + | + | + | + | 20 | 2.0 |
| 13 | + | + |  | + | + | + | + |  | + |  | 19 | 1.9 |
| 14 |  | + |  | + | + |  | + | + | + |  | 17 | 1.7 |
| 15 |  | + |  | + |  |  |  |  | + | + | 16 | 1.6 |
| 16 |  | + |  | + |  |  | + |  | + |  | 16 | 1.6 |
| 17 |  | + |  | + |  |  | + | + | + |  | 16 | 1.6 |
| 18 |  | + |  | + | + |  | + |  | + |  | 16 | 1.6 |
| 19 |  | + |  | + | + | + |  | + | + |  | 16 | 1.6 |
| 20 |  | + |  | + |  | + | + | + | + |  | 15 | 1.5 |

*\*Variable is kept as non-linear effect using natural cubic splines with three knots*
*Selected model is ranked first (bold face)*

**Table S8.** Model stability for adults.

| Predictors | Initial model |  | Selected model |  | Bootstrap inclusion frequency (%) | RMSD ratio | Relative conditional bias (%) | Bootstrap median | Bootstrap 2.5<sup>th</sup> percentile | Bootstrap 97.5<sup>th</sup> percentile |
| --- | --- | --- | --- | --- | --- | --- | --- | --- | --- | --- |
|  | Estimate | Standard error | Estimate | Standard error |  |  |  |  |  |  |
| (Intercept) | -16.6085 | 3.2632 | -16.6780 | 3.2475 | 100.0 | 1.3263 | 2.0586 | -16.7262 | -26.4755 | -9.8095 |
| SDC-1 | 1.1524 | 0.2708 | 1.1740 | 0.2607 | 99.2 | 1.2745 | 3.0018 | 1.1767 | 0.5450 | 1.8615 |
| IL-8 | 0.5473 | 0.1427 | 0.5544 | 0.1411 | 98.9 | 1.2973 | 5.8880 | 0.5739 | 0.2490 | 0.9520 |
| Ferritin | -0.2785 | 0.0916 | -0.2682 | 0.0883 | 94.6 | 1.2479 | 7.9330 | -0.2845 | -0.5012 | 0 |
| sTREM-1 | -0.2961 | 0.1499 | -0.2864 | 0.1492 | 66.5 | 1.4191 | 28.1687 | -0.2807 | -0.6448 | 0 |
| IL-1RA | 0.2582 | 0.1583 | 0.2557 | 0.1427 | 62.3 | 1.4788 | 54.9829 | 0.2494 | 0 | 0.7255 |
| IP-10 (ns1)* | -1.4427 | 1.0592 | -0.8269 | 0.6118 | 59.8 | 1.6064 | 36.8995 | -0.2108 | -5.2628 | 0.7003 |
| IP-10 (ns2)* | -0.1027 | 0.5763 | 0.1139 | 0.4976 | 59.8 | 1.2060 | 43.1473 | 0 | -1.7021 | 1.2589 |
| sCD163 | 0.2056 | 0.1287 | 0.2351 | 0.1253 | 59.1 | 1.3333 | 51.3932 | 0.2148 | 0 | 0.4989 |
| CRP | 0.0863 | 0.0990 |  |  | 36.3 | 1.1203 | 129.8186 | 0 | 0 | 0.3048 |
| VCAM-1 | 0.0660 | 0.0685 |  |  | 35.4 | 1.3808 | 98.3111 | 0 | -0.0923 | 0.2714 |
| Ang-2 | -0.0246 | 0.1193 |  |  | 21.8 | 1.0047 | 62.8642 | 0 | -0.2792 | 0.3008 |

\*As IP-10 is kept as non-linear effect using natural cubic splines with 3 knots, there are two terms of this variable in the model
RMSD: root mean squared difference

## Appendix 7. Results of variable selection using different approaches

**Table S9.** Results of variable selection for children.

|  | VCAM-1 | SDC-1 | Ang-2 | IL-8 | IP-10 | IL-1RA | sCD163 | sTREM-1 | Ferritin | CRP |
| --- | --- | --- | --- | --- | --- | --- | --- | --- | --- | --- |
| Best subset |  | + | + | + | + | + |  |  | + |  |
| Backward elimination |  | + | + | + | + | + |  |  | + |  |
| Forward selection |  | + | + | + | + | + |  |  | + |  |
| Stepwise forward |  | + | + | + | + | + |  |  | + |  |
| Stepwise backward |  | + | + | + | + | + |  |  | + |  |
| Augmented backward elimination | + | + | + | + | + | + |  |  | + |  |
| Bayesian projection |  |  | + | + | + | + |  |  | + |  |

**Table S10.** Results of variable selection for adults.

|  | VCAM-1 | SDC-1 | Ang-2 | IL-8 | IP-10* | IL-1RA | sCD163 | sTREM-1 | Ferritin | CRP |
| --- | --- | --- | --- | --- | --- | --- | --- | --- | --- | --- |
| Best subset |  | + |  | + | + | + | + | + | + |  |
| Backward elimination |  | + |  | + | + | + | + | + | + |  |
| Forward selection |  | + |  | + |  | + |  | + | + |  |
| Stepwise forward |  | + |  | + |  | + |  | + | + |  |
| Stepwise backward |  | + |  | + | + | + | + | + | + |  |
| Augmented backward elimination | + | + |  | + | + | + | + | + | + |  |
| Bayesian projection |  | + |  | + |  |  |  |  |  |  |

\*Variable is kept as non-linear effect using natural cubic splines with 3 knots

## Appendix 8. STROBE Statement - Checklist of items

**Title:** Combination of inflammatory and vascular markers in the febrile phase of dengue is associated with more severe outcomes

|  | Item No | Recommendation | Comments or page(s) reported |
| --- | --- | --- | --- |
| Title and abstract | 1 | (a) Indicate the study’s design with a commonly used term in the title or the abstract | 1 |
|  |  | (b) Provide in the abstract an informative and balanced summary of what was done and what was found | 4 |
| Introduction |  |  |  |
| Background/rationale | 2 | Explain the scientific background and rationale for the investigation being reported | 5-6 |
| Objectives | 3 | State specific objectives, including any prespecified hypotheses | 6 |
| Methods |  |  |  |
| Study design | 4 | Present key elements of study design early in the paper | 7 |
| Setting | 5 | Describe the setting, locations, and relevant dates, including periods of recruitment, exposure, follow-up, and data collection | 7 |
| Participants | 6 | (a) Give the eligibility criteria, and the sources and methods of selection of participants. Describe methods of follow-up | 7-8 |
|  |  | (b) For matched studies, give matching criteria and number of exposed and unexposed | Not applicable |
| Variables | 7 | Clearly define all outcomes, exposures, predictors, potential confounders, and effect modifiers. Give diagnostic criteria, if applicable | 7-9, Appendices 1-2 |
| Data sources/<br>measurement | 8* | For each variable of interest, give sources of data and details of methods of assessment (measurement). Describe comparability of assessment methods if there is more than one group | 7, Appendices 1-2 |
| Bias | 9 | Describe any efforts to address potential sources of bias | 7 |
| Study size | 10 | Explain how the study size was arrived at | 7-8, Figure 1 |
| Quantitative variables | 11 | Explain how quantitative variables were handled in the analyses. If applicable, describe which groupings were chosen and why | 9, Appendix 3 |
| Statistical methods | 12 | (a) Describe all statistical methods, including those used to control for confounding | 9, Appendix 3 |
|  |  | (b) Describe any methods used to examine subgroups and interactions | 9, Appendix 3 |
|  |  | (c) Explain how missing data were addressed | Not applicable |
|  |  | (d) If applicable, explain how loss to follow-up was addressed | Not applicable |
|  |  | (e) Describe any sensitivity analyses | Appendix 3 |
| Results |  |  |  |
| Participants | 13* | (a) Report numbers of individuals at each stage of study—eg numbers potentially eligible, examined for eligibility, confirmed eligible, included in the study, completing follow-up, and analysed | 10, Figure 1 |
|  |  | (b) Give reasons for non-participation at each stage | Figure 1 |
|  |  | (c) Consider use of a flow diagram | Figure 1 |
| Descriptive data | 14* | (a) Give characteristics of study participants (eg demographic, clinical, social) and information on exposures and potential confounders | 10-11, Table 1 |
|  |  | (b) Indicate number of participants with missing data for each variable of interest | Tables 1-2 |
|  |  | (c) Summarise follow-up time (eg, average and total amount) | Not applicable |
| Outcome data | 15* | Report numbers of outcome events or summary measures over time | 10-11, Table 1 |
| Main results | 16 | (a) Give unadjusted estimates and, if applicable, confounder-adjusted estimates and their precision (eg, 95% confidence interval). Make clear which confounders were adjusted for and why they were included | Figure 3, S3-S5, Tables 3, S2-S4 |
|  |  | (b) Report category boundaries when continuous variables were categorized | Not applicable |
|  |  | (c) If relevant, consider translating estimates of relative risk into absolute risk for a meaningful time period | Not applicable |

|  | **Item**<br>**No** | **Recommendation** | **Comments or page(s) reported** |
| --- | --- | --- | --- |
| Other analyses | 17 | Report other analyses done—eg analyses of subgroups and interactions, and sensitivity analyses | Tables 4-5, S5-S10 |
| **Discussion** |  |  |  |
| Key results | 18 | Summarise key results with reference to study objectives | 12 |
| Limitations | 19 | Discuss limitations of the study, taking into account sources of potential bias or imprecision. Discuss both direction and magnitude of any potential bias | 14 |
| Interpretation | 20 | Give a cautious overall interpretation of results considering objectives, limitations, multiplicity of analyses, results from similar studies, and other relevant evidence | 12-14 |
| Generalisability | 21 | Discuss the generalisability (external validity) of the study results | 12-14 |
| **Other information** |  |  |  |
| Funding | 22 | Give the source of funding and the role of the funders for the present study and, if applicable, for the original study on which the present article is based | 16 |

\*Give information separately for exposed and unexposed groups.
**Note:** An Explanation and Elaboration article discusses each checklist item and gives methodological background and published examples of transparent reporting. The STROBE checklist is best used in conjunction with this article (freely available on the Web sites of PLoS Medicine at http://www.plosmedicine.org/, Annals of Internal Medicine at http://www.annals.org/, and Epidemiology at http://www.epidem.com/). Information on the STROBE Initiative is available at http://www.strobe-statement.org.

